# Orthogonal Functions for Evaluating Social Distancing Impact on CoVID-19 Spread

**DOI:** 10.1101/2020.06.30.20143149

**Authors:** Genghmun Eng

## Abstract

Early CoVID-19 growth often obeys: 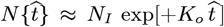, with *K*_*o*_ = [(ln 2)/(*t*_*dbl*_)], where *t*_*dbl*_ is the pandemic *doubling time*, prior to society-wide *Social Distancing*. Previously, we modeled *Social Distancing* with *t*_*dbl*_ as a linear function of time, where *N* [*t*] **1** ≈ exp[+*K*_*A*_ *t*/ (1+,*γ*_*o*_*t*)] is used here. Additional parameters besides {*K*_*o*_, *γ*_*o*_} are needed to better model different *ρ*[*t*] = *dN* [*t*]/*dt* shapes. Thus, a new *Orthogonal Function Model* [*OFM*] is developed here using these orthogonal function series:

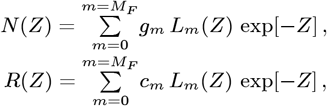

where *N* (*Z*) and *Z*[*t*] form an implicit *N* [*t*] *N* (*Z*[*t*]) function, giving:

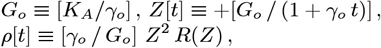

with *L*_*m*_(*Z*) being the *Laguerre Polynomials*. At large *M*_*F*_ values, nearly *arbitrary* functions for *N* [*t*] and *ρ* [*t*] = *dN* [*t*]/*dt* can be accommodated. How to determine {*K*_*A*_, *γ*_*o*_} and the {*g*_*m*_; *m* = (0, +*M*_*F*_)} constants from any given *N* (*Z*) dataset is derived, with *ρ* [*t*] set by:

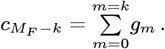

The *bing com* USA CoVID-19 data was analyzed using *M*_*F*_ = (0, 1, 2) in the *OFM*. All results agreed to within about 10 percent, showing model robustness. Averaging over all these predictions gives the following overall estimates for the number of USA CoVID-19 cases at the pandemic end:

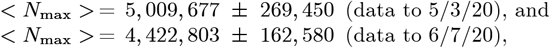

which compares the pre- and post-early May *bing com* revisions. The CoVID-19 pandemic in Italy was examined next. The *M*_*F*_ = 2 limit was inadequate to model the Italy *ρ* [*t*] pandemic tail. Thus, regions with a quick CoVID-19 pandemic shutoff may have additional *Social Distancing* factors operating, beyond what can be easily modeled by just progressively lengthening pandemic *doubling times* (with *13 Figures*).

## 1 Introduction

The early stages of the CoVID-19 coronavirus pandemic around the world showed a nearly exponential rise in the number of infections with time. If a significant fraction of the population gets infected (*“saturated pandemic”*), exponential growth no longer applies. However, *Social Distancing* can also mitigate exponential growth, enabling pandemic shutoff with only a small fraction of the population being infected (*“dilute pandemic”*). Let 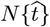 be the total number of CoVID-19 cases in any given region, with 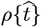 being the predicted number of daily new CoVID-19 cases, so that:

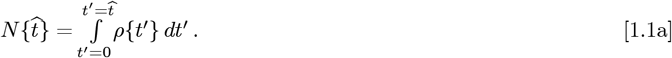

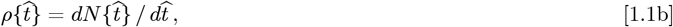

On 3/25/2020, the *Institute of Health Metrics and Evaluation, University of Washington* (IHME) released their initial model for CoVID-19 spread^**1**^ where:

> “*The cumulative death rate for each location is assumed*
>
> *to follow a parametrized Gaussian error function* “

Since the IHME 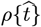 used Gaussians, their projections assumed that the rise to the pandemic peak and its subsequent fall would be symmetric. Their implicit assumption was that the amount of *Social Distancing* was exactly what was needed to make their model predictions true. Given a sharp 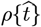 rise, our concern was that the IHME model did not allow 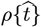 to decrease gradually.

As a result, we developed an alternative CoVID-19 spread model, which assumed^2^ *Social Distancing* gradually lengthens the CoVID-19 *doubling time*. The initial exponential growth factor *K*_*o*_ = [(ln 2)/*t*_*dbl*_] was used as a starting point, where *t*_*dbl*_ was the initial *doubling time*. A new *Social Mitigation Parameter* [*SMP*] *a*_*S*_ was introduced to account for society-wide *Social Distancing* measures. A linear function was used for *doubling time* lengthening as a simple extension beyond a constant *K*_*o*_, giving:

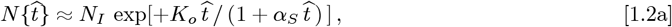

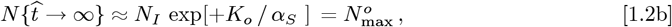

as an *Initial Model*^**1**^ for the number of CoVID-19 cases, where 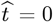 was the start of society-wide *Social Distancing*. Both 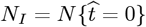 and 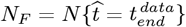, as the most recently available data, were treated as fixed points. A minimum *root-mean-square* (*rms*) error datafit, using a logarithmic Y-axis, sets the {*K*_*o*_, *a*_*S*_} values. The resulting 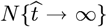 of Eq. [1.2b] is the predicted final number of CoVID-19 cases at the pandemic end.

On 4/29/2020, we sent our preprint^**1**^ to the IHME, the Los Angeles Department of Public Health (LADPH), and to Profs. Goldenfeld and Maslov at UOI (University of Illinois at Urbana-Champaign), who were preparing a 2-day nationwide CoVID-19 remote-learning seminar for 5/6/2020 and 5/8/2020.

Also, on 4/29/2020, the IHME electronically published their 12^*th*^ CoVID-19 update, using their 3/25/2020 model. A graphic display of their most recent 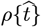 projections showed a symmetric rise and fall. This graph was widely publicized by Dr. Alan Boyle, who was following the IHME work, summarizing it for general audiences^**3**–**5**^. Since our Eqs. [1.2a]-[1.2b] model gave substantially different 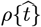 predictions than IHME, we added a note to that effect in our pre-print, submitting the final pre-print to MedRxiv on 5/4/2020, where it was accepted and published on-line on 5/8/2020.

Concurrently, on 5/4/2020, IHME published their 13^*th*^ CoVID-19 update^**6**^, where everything changed. Dr. Alan Boyle^**5**^ summarized those changes with a note that: “[*IHME*] *researchers acknowledged that their previous modeling wasn’t sophisticated enough*” Both IHME graphical predictions for 4/29/2020 and 5/4/2020 are shown in **Figure 1**, to highlight this change.

**Figure 1:**
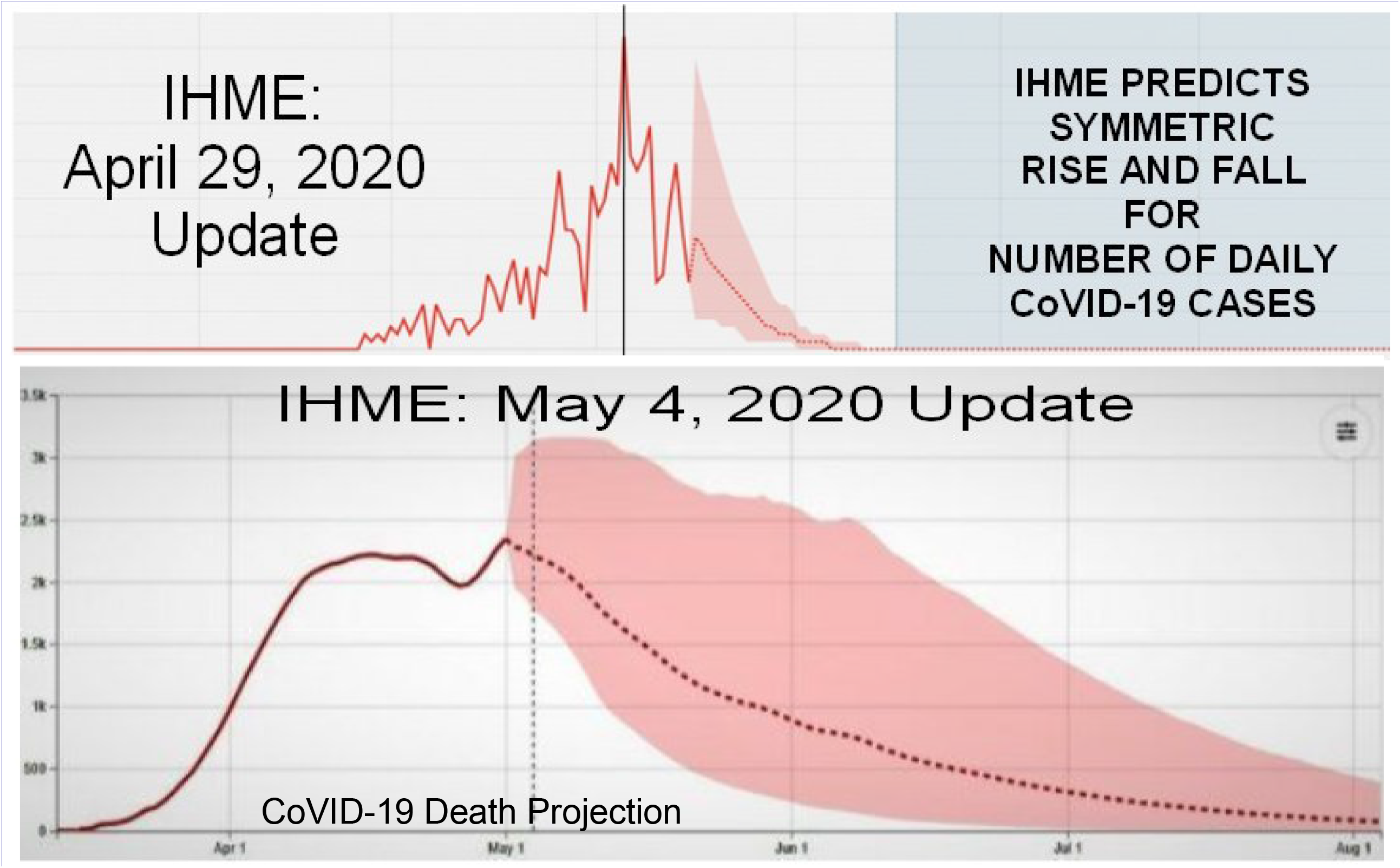
Comparison of IHME CoVID-19 Projections, 29 April 2020 vs 4 May 2020. CDC CoVID-19 Website highlighted IHME Projections prior to the IHME May 2020 update.

On 5/6/2020 and 5/8/2020, Profs. Goldenfeld and Maslov presented their UOI team’s supercomputer-based 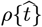 CoVID-19 projections, which also were very asymmetric. Although mathematical details for the UOI and new IHME projections are not known, virtually all 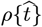 CoVID-19 projections are now asymmetric, as the developing CoVID-19 data also appears to be.

Since our *Initial Model* had only two data fitting parameters {*K*_*o*_, *a*_*S*_}, we became concerned that those two parameters might not be sufficient to adequately describe all the different 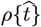 shapes observed. To correct this potential defect, a new *Orthogonal Function Model* [*OFM*] is developed here to allow more accurate descriptions for a variety of 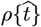 shapes, using additional mathematical techniques derived herein. This *OFM* extends Eqs. [1.2a]-[1.2b], and provides additional fitting parameters to improve 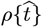 projections.

## 2. *Orthogonal Function Model [OFM]* Elements

The following items and methods were developed as part of this *OFM* to improve CoVID-19 projections for a variety of 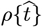 data shapes.

**First**, the 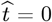 point in Eq. [1.2a] was time-shifted so that *N* [*t* = 0] ≡ **1**. This *t* = 0 point now provides an estimate for the CoVID-19 pandemic starting point, replacing the above 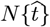 with this time-shifted version:

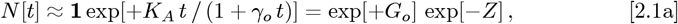

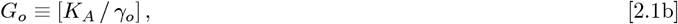

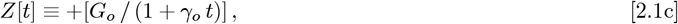

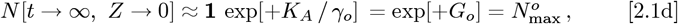

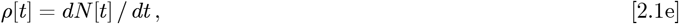

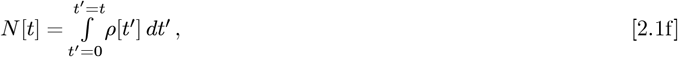

which enables Eq. [2.1a] to become a 1-term approximation for a larger function series. Actual data provides the {*N*_*I*_, *N*_*F*_} values. However, these {*t* = *t*_*I*_, *N* [*t*_*I*_] ≡ *N*_*I*_} and {*t* = *t*_*F*_, *N* [*t*_*F*_] ≡ *N*_*F*_} limits are now used to set {*K*_*A*_, *G*_*o*_, *γ*_*o*_} > 0, so that the 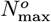 of Eq. [1.2b] and Eq. [2.1d] match exactly.

**Second**, when *Z* → 0 in Eq. [2.1c] then *t* → ∞; while *Z* → + ∞ corresponds to *t* → (− 1/,*γ*_*o*_)+ *ε*, where *ε* is arbitrarily small and positive. Since *N* [*t* = 0] = **1**, the *t* < 0domain has *N* [*t*] < 1, while setting a particular time as the *N* [*t*] = 0 point. Since the **1** > *N* [*t*] > 0 regime has no impact on this overall analysis, virtually any decreasing function tail for the *Z* → + ∞ limit should be allowed.

**Third**, instead of generalizing Eq. [2.1a] using time, it is easier to use functions of *Z*, where *Z* is given by Eq. [2.1c]. It results in these *N* (*Z*) and *R*(*Z*) substitutes for *N* [*t*] and *p*[*t*]:

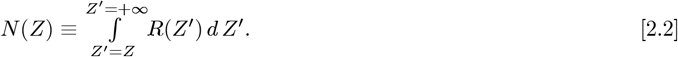

Given explicit functions of *Z*, both *N* (*Z*) and *R*(*Z*) in Eq. [2.2] go from large − *Z* to smaller − *Z* values at longer times, eventually approaching the *Z* = 0 point. Together, *N* (*Z*) and *Z*[*t*] create an implicit *N* (*Z*[*t*]) *N* [*t*] function, and *R*(*Z*) and *Z*[*t*] create another implicit *R*(*Z*[*t*]) *R*[*t*] function. A standard change of variables converts them back into being explicit functions of time:

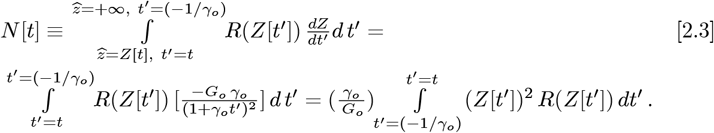

It gives these equivalences between and Eq. [2.2] and Eq. [2.1f]:

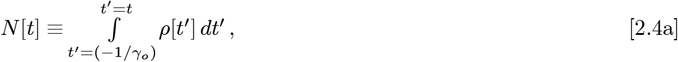

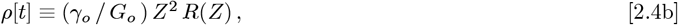

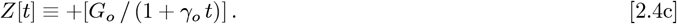

**Fourth**, to allow additional data fitting parameters, the *OFM* replaces the 1-term approximation of Eq. [2.1a] with these orthogonal function series:

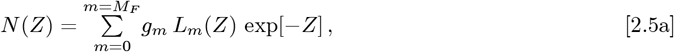

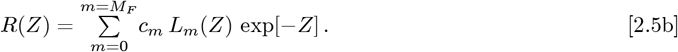

These series have exp[−*Z*] as their weighting function, while keeping the Eqs. [2.1b]-[2.1c] definitions for {*Z, G*_*o*_, *γ*_*o*_}. The {*g*_*m*_; *m* = (0, +*M*_*F*_)} and {*c*_*m*_; *m* = (0, +*M*_*F*_)} coefficients are constants that can be derived from each dataset. For a wide range of *N* (*Z*) and *R*(*Z*) functions, larger *M*_*F*_ values and more {*L*_*m*_(*Z*); *m* = (0, +*M*_*F*_)} terms give progressively better matches to practically any *arbitrary function*. This feature is what enables improved datafits over a variety of measured *N* [*t*] and *p*[*t*] curves.

**Fifth**, the *OFM* uses the {*N*_*I*_ [*t*_*I*_], *N*_*F*_ [*t*_*F*_]}data end-points to set {*G*_*o*_,, *γ*_*o*_} in Eq. [2.4c], and define *Z*, allowing the *OFM* to provide best fits over the whole data range of *Z* or *t*, while these end points are fixed in the *Initial Model*.

The difference between: (a) using the whole data range for fitting, versus (b) using the data end points for fitting, is most evident when comparing Eq. [2.1a] to Eq. [2.5a]. In Eq. [2.1a], *N* [*t*] = *G*_*o*_ exp[−*Z*] where *G*_*o*_ has a pre-set value, whereas in Eq. [2.5a], *N* (*Z*) = *g*_0_ exp[−*Z*] for *M*_*F*_ = 0, the *g*_0_ parameter is determined by fitting over the whole data range.

**Sixth**, both *Z* and *t* essentially span from {0 + ∞}. Using exp[−*Z*] as a weighting function over that domain makes the choice of *L*_*m*_(*Z*) in Eq. [2.5a]- [2.5b] unique. They are the *Laguerre Polynomials*, with the first few being:

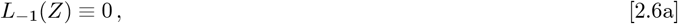

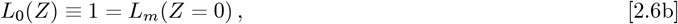

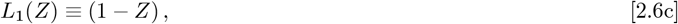

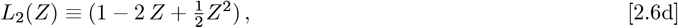

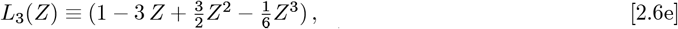

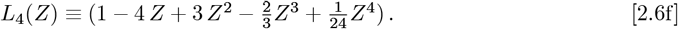

Some important properties of the *Laguerre Polynomials* are:

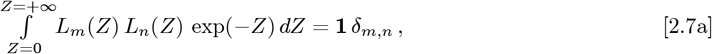

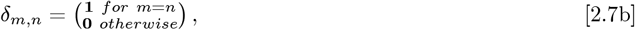

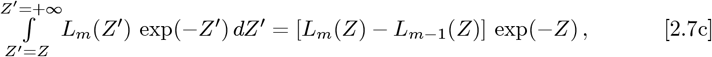

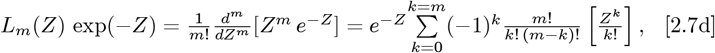

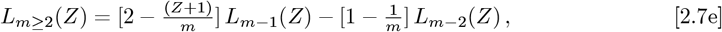

where Eq. [2.7a] defines an *orthogonal function set*. Given **n** is an integer in Eq. [2.7d], **n**-*factorial* (**n**!) is defined as the product:

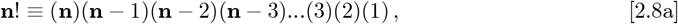

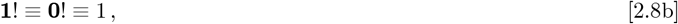

along with *factorials* involving negative integers not being allowed.

**Seventh**, when data are used to determine the {*g*_*m*_; *m* = (0, *M*_*F*_)} constants for the Eq. [2.5a] *N* (*Z*) analytic approximation, an equivalently precise *R*(*Z*) is set by Eq. [2.2] and Eq. [2.5b], with its {*c*_*m*_; *m* = (0, *M*_*F*_)} constants being:

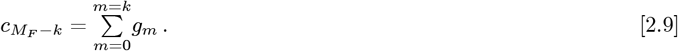

This simple form of Eq. [2.9] arises from the fact that *L*_*m*_(*Z* =) = 1. Also, Eq. [2.5a] and Eq. [2.9] combine to give:

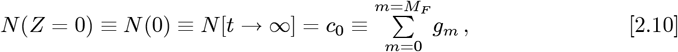

as a new predicted total number of CoVID-19 cases at pandemic end.

**Eighth**, the {*g*_*m*_; *m* = (0, *M*_*F*_)} constants can be arranged in a 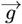 form, with comparable constants for *R*(*Z*) from Eq. [2.2] arranged in a 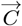 form. It allows Eq. [2.9] to be written as:

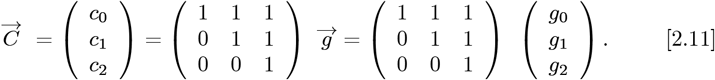

Once the {*g*_*m*_; *m* = (0, *M*_*F*_)} constants are found, the *c*_0_ value in Eq. [2.11] becomes the {*M*_*F*_ + 1}-term replacement value for the predicted total number of CoVID-19 cases at the pandemic end, which refines the initial 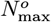 value of Eq. [1.2b] or Eq. [2.1d]. How to determine {*K*_*A*_, *G*_*o*_, _*o*_} and the {*g*_*m*_; *m* = (, *M*_*F*_)} constants in Eq. [2.5a] from a given set of data, is derived next.

## 3. Finding {*K*_*A*_, *γ*_*o*_} for *Z*[*t*] from Data

For a given dataset, the *OFM* begins with using Eq. [1.2a] to set {*K*_*o*_, *α*_*S*_}, as in our *Initial Model*. Society-wide *Social Distancing* is assumed to occur at or before the time *t*_*I*_, where *N*_*I*_ cases are already observed. Since the most recently available data at *t*_*F*_ has *N*_*F*_ cases, Eq. [2.1a] becomes:

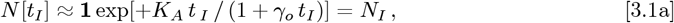

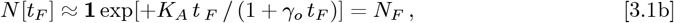

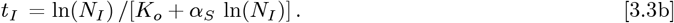

which using the new *t* = point for the *OFM*. Evaluating *N* [*t* < *t*_*I*_] for *t*< *t*_*I*_ estimates what the pandemic prior history might have been, had society-wide *Social Distancing* already been in place. Evaluating *N* [*t*> *t*_*F*_] for *t*> *t*_*F*_ estimates how the pandemic evolves assuming these *Social Distancing* measures remain in place. The prior Eq. [1.2a] gave:

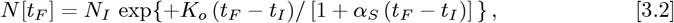

with the *t*_*F*_ limit of Eq. [3.2] giving:

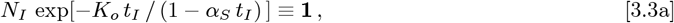

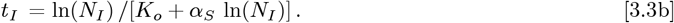

Here, Eq. [3.3b] sets the precise *t*_*I*_ time shift needed to convert from Eq. [1.2a] to Eq. [2.1a], which is easier to generalize. In addition, the *t* = point of Eq. [2.1a] gives *N* [*t* → 0] = **1** as an estimate for the pandemic starting point. Since *t*_*I*_ and (*t*_*F*_ − *t*_*I*_) sets *t*_*F*_, the Eqs. [3.1a]-[3.1b] fully determine {*K*_*A*_, *γ*_*o*_}, without needing any recalculations on the original dataset. Taking various ratios of Eq. [3.1b] to Eq. [3.1a] gives:

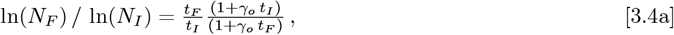

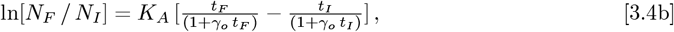

as separable equations to first find *γ*_*o*_, then *K*_*A*_, with these results:

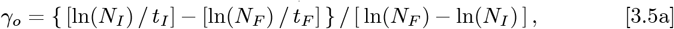

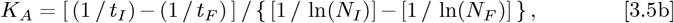

which sets the *Z*[*t*] function in Eq. [2.1c] or Eq. [2.4c].

## 4. Determining the g_*m*_ Constants from Data

When data for *N*_*data*_(*Z*) are given over the whole *Z* = {0^+^,∞^−^} range, the *g*_*n*_ constants for Eq. [2.5a] are exactly determined via:

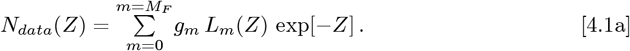

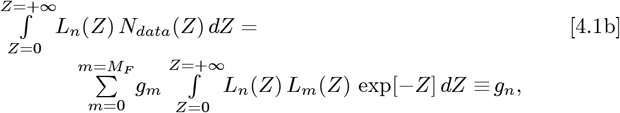

where the *Laguerre Polynomial* orthogonality condition of Eq. [2.7a] forces the Eq. [4.1b] sum to reduce to one term.

When the *N*_*data*_(*Z*) only spans a finite range of: *t*_*I*_ <*t*< *t*_*F*_ and *Z*_min_ <*Z* < *Z*_max_, an extrapolation of *N*_*data*_(*Z*) for (*Z* < *Z*_min_) and (*Z* > *Z*_max_) is needed. One method could set *N*_*data*_(*Z* < *Z*_min_) and *N*_*data*_(*Z* > *Z*_max_), which results in these Eqs. [4.1a]-[4.1b] cognates:

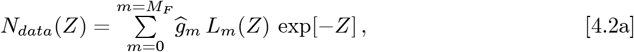

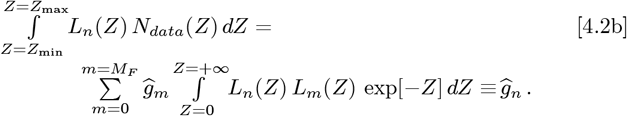

Its advantages are (a) for m ≠ n every *ĝ*_*m*_ and *ĝ*_*n*_ are independent, as in orthogonal functions; and (b) these *ĝ*_*m*_ values provide new estimates for the *Ndata*(*Z* < *Z*_max_) and *Ndata*(*Z* > *Z*min) regimes. But since *N*_*data*_(*Z* < *Z*_min_) and *N*_*data*_(*Z* > *Z*_max_) were originally assumed to vanish, this method is inconsistent. Alternatively, adding reasonable “*tails*” to the data could extend the original *N*_*data*_(*Z*) domain, but those functions are not always known.

The third path, used here, takes the Eq. [4.1a] “*final answer* “as a *self-consistent* extrapolation for (*Z* < *Z*_min_) and (*Z* > *Z*_max_), while retaining the *N*_*data*_(*Z*) values for the (*Z*_max_ ≥ *Z* ≥ *Z*_min_) regime. It replaces Eq. [4.1b] with:

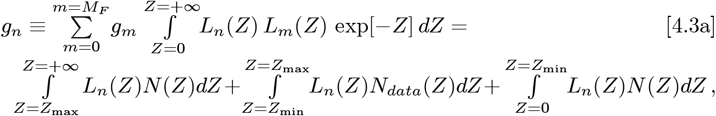

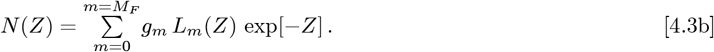

The {*g*_*m*_; *m* = (0, *M*_*F*_)} now appears on both sides of each Eq. [4.3a] *g*_*n*_-equation, which is handled as follows. Defining:

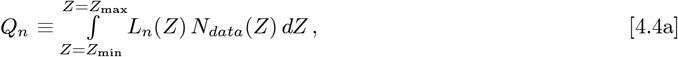

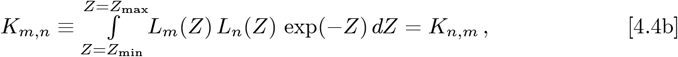

Eqs. [4.3a]-[4.3b] can be re-written as a 3 x 3 matrix <u>**M**</u>_3_, which relates a data-driven 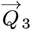 vector to a resultant 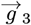 vector:

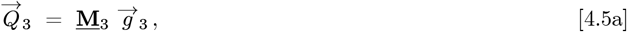

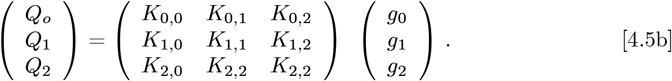

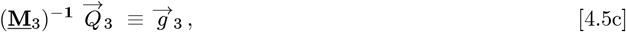

where (<u>**M**</u>_3_)^-**1**^ is the matrix inverse of <u>**M**</u>_3_. hen {*Z*_min_, *Z*_max_} → {0, +∞}, this <u>**M**</u>_3_ becomes the Identity Matrix. The following *k*_*m,n*_(*Z*) integrals set *K*_*m,n*_:

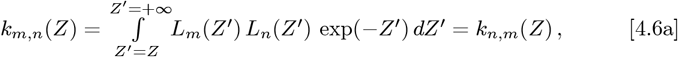

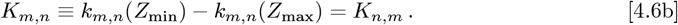

The *k*_*m,n*_(*Z*) integrals can be determined using Eq. [2.7c], which gives:

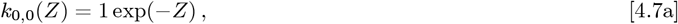

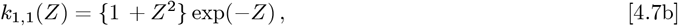

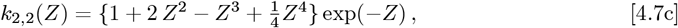

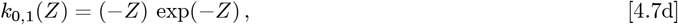

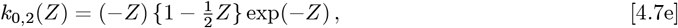

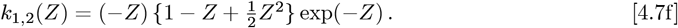

To extract {*g*_0_, *g*_1_, *g*_2_}, the 3 x 3 symmetric <u>**M**</u>_3_ matrix needs inversion:

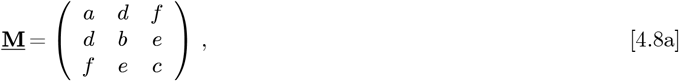

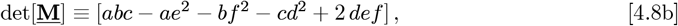

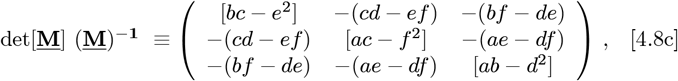

which determines {*g*_0_, *g*_*1*_, *g*_*2*_} from the {*Q*_0_, *Q*_*1*_, *Q*_*2*_} data. A best fit *N* (*Z*) for *Z* = {0^+^, ∞^−^} results, along with an equivalent fit for *R*(*Z*) using Eq. [2.9]. Instead of having to find the best {*g*_0_, *g*_1_, *g*_2_} triplet, one could find the best 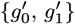 by just using using {*Q*_0_, *Q*_1_} and an <u>**M**</u>_2_ sub-matrix; or one could find the best 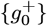 by itself by just using {*Q*_0_} and an <u>**M**</u>_1_ sub-matrix:

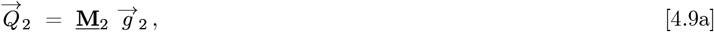

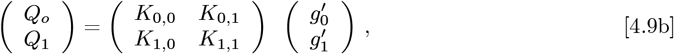

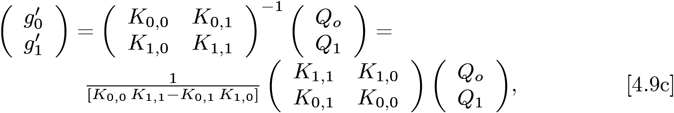

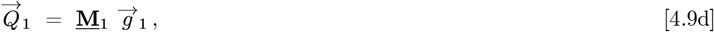

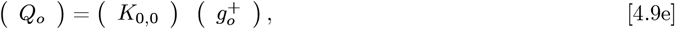

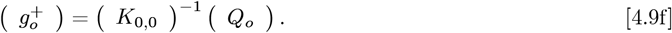

When the *N*_*data*_(*Z*) is comprised of *j* = {1, 2,*…J*} discrete values between {*Z*_min_, *Z*_max_}, with each *Z*_*j*_ having an 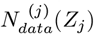 value, the Eq. [4.4a] integral needs to be replaced by a sum. Let:

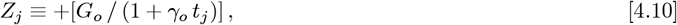

with *Z*_0_ = *Z*_1_and *Z*_*J + 1*_ = *Z*_*J*_, the *Q*_*n*_ replacement for Eq. [4.4a] is:

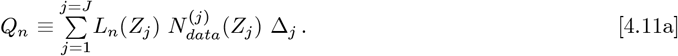

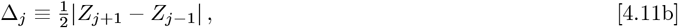

with the *N* [*t*] and *ρ* [*t*] being set by Eq. [2.3] and Eqs. [2.4a]-[2.4c].

Finally, the Eqs. [4.7a]-[4.7f] *k*_*m,n*_(*Z*) integrals are easy to compute for 0 ≤*m* ≤2 and 0 ≤*n* ≤ 2. But the general case is not well-known or tabulated in many *Tables of Integrals*. The key is how to express a product of two *Laguerre Polynomials* efficiently as a sum over a larger set of single *Laguerre Polynomials*, so as to convert the Eq. [4.6a] integrals into the Eq. [2.7c] form.

This problem was originally solved by G. N. Watson^**7**^ in 1938, and simplified by J. Gillis and G. Weiss^**8**^ in 1960. It is a sum of terms, where each coefficient contains four different *factorials* involving integers. Their key result is:

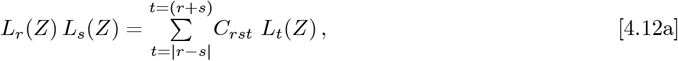

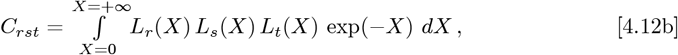

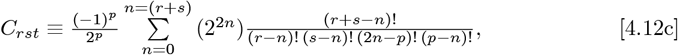

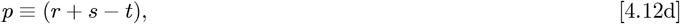

where ALL terms in the sum for *n* = {0, (*r* + *s*)} also have an implicit requirement that none of the integer arguments for any of the *factorials* can be negative. Thus, all terms with negative arguments for the *factorial* must be omitted. Nowadays, this calculation can be done on a computer, but it would have been difficult in 1960, and nearly impossible in 1938.

## 5 USA: *Orthogonal Function Model* Results

This USA analysis only uses data after mid-March 2020, when several State Governors instituted mandatory *Mitigation Measures*. The widely available *bing com* CoVID-19 data^**1**^ for the USA had these limits:

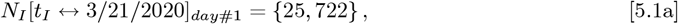

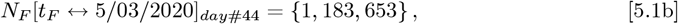

with (*t*_*F*_ − *t*_*I*_) = 43 *days*. Our *Initial Model* of Eq. [1.2a] sets these parameter values for the USA:

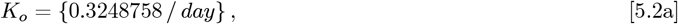

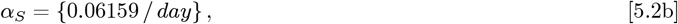

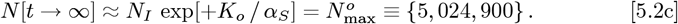

Using Eq. [3.3b] for *t*_*I*_ and *t*_*F*_ sets:

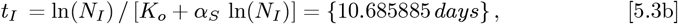

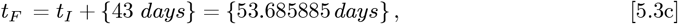

for use in the *OFM*. **Figures 2**-**3** show how this *Initial Model*, by itself, compares to the USA CoVID-19 data. **Figure 2** uses a logarithmic Y-axis for the predicted total number of CoVID-19 cases, and **Figure 3** shows the daily new CoVID-19 case predictions on a linear Y-axis plot.

**Figure 2:**
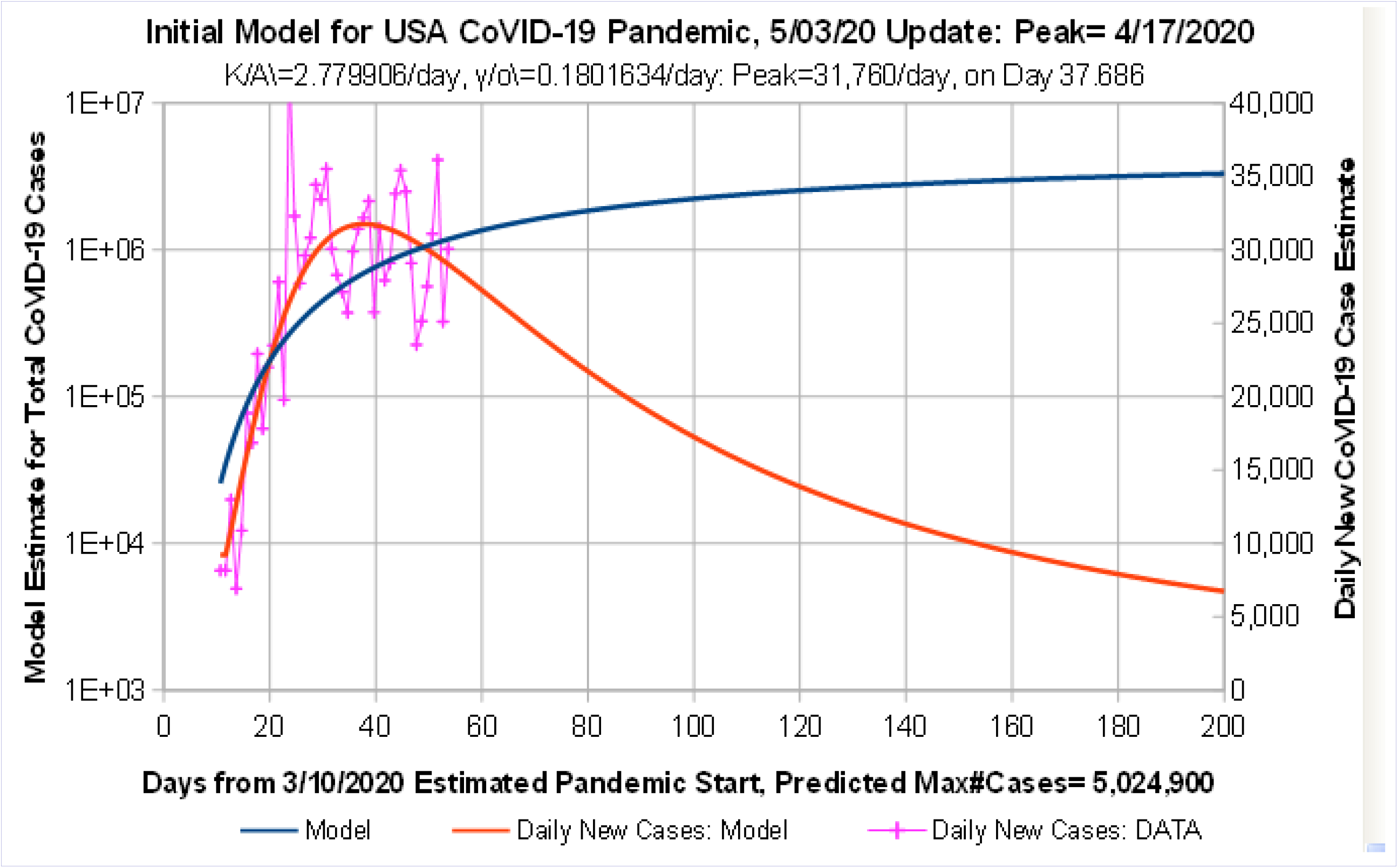
*Initial Model* for USA CoVID-19 Projections using data up to 5/3/2020. Predicted Number of Daily CoVID-19 Cases has a peak of 31,760 cases/day on 4/17; with 5,024,900 cases total; and ∼6,757 new cases/day at Day 200 on 9/26/2020.

**Figure 3:**
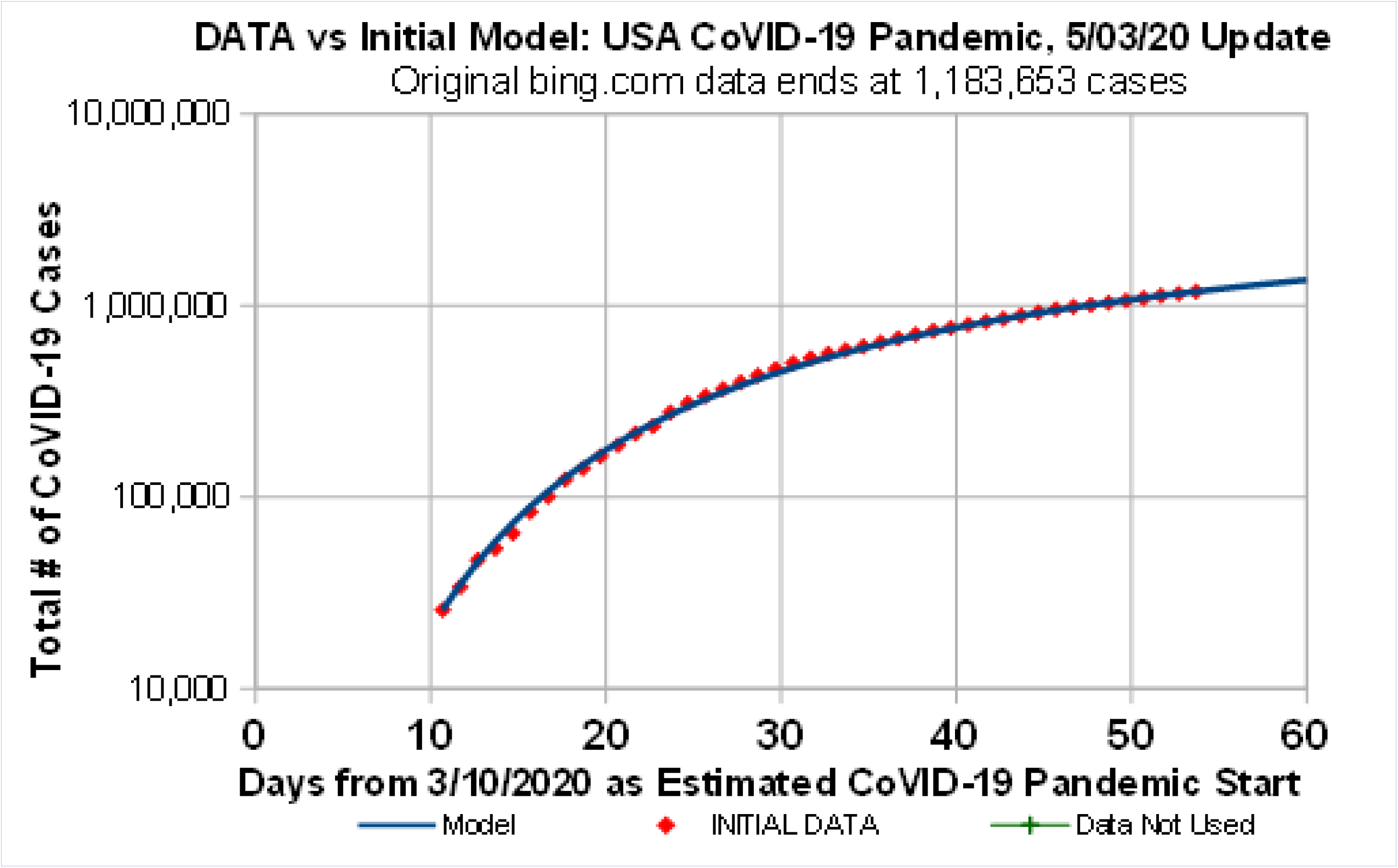
*Initial Model* for USA CoVID-19 Projections vs data up to 5/3/2020. Original *bing.com* data up to 5/3/2020 are shown, prior to their *new reporting method*. Data starts slightly below, then goes slightly above the *Initial Model* prediction line.

The daily new case data exhibits large day-to-day variations, likely due to reporting delays, among other factors. This *Initial Model* for the USA has a predicted maximum of ~31, 760 new cases per day at Day 37.686 on 4/17/2020, along with ~6, 757 new cases per day still occurring at Day 2 on 9/26/2020.

The time axis in **Figure 2** is different than in our previous paper^**2**^, due to the time shift of Eq. [2.1a], where the new *t* = point estimates the CoVID-19 pandemic starting point being on 3/10/2020. Even if *Social Distancing* had been in effect at the start of the pandemic, **Figure 2** shows that the *N*_*I*_ [*t*_*I*_] = {25, 722} level still could have been reached in 10 – 11 days.

**Figures 3** compares the measured data for the total number of CoVID-19 cases after *Social Distancing* started, to the early-time portion of this *Initial Model*. That comparison shows that the early-time data starts off a little below the curve; the later-time data rises a bit above the curve; and the final-time data again matches the curve, since it is a fixed point for this analysis.

These predictions assume: (I) The present *Mitigation Measures* are continued; (II) No “*second wave*” of infection or re-infection occurs; and (III) No further *Mitigation Measures* are taken to reduce the number of CoVID-19 cases.

These *Initial Model* results are first refined by applying the Eq. [2.1a] time shift, with Eqs. [3.5a]-[3.5b] setting these {*γ*_*o*_, *K*_*A*_, *G*_0_} values:

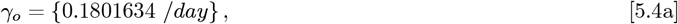

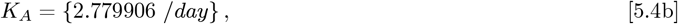

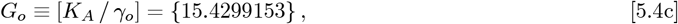

where Eq. [3.1c] also gives:

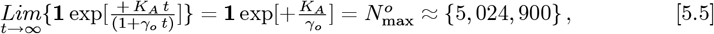

which matches Eq. [5.2c], as it should. Then:

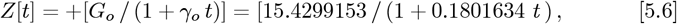

defines *Z* for the *OFM*, where:

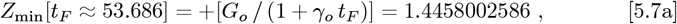

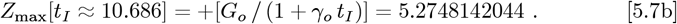

The resultant Eq. [4.5b] <u>**M**</u>_3_ matrix of *K*_*m*,*n*_ entries is:

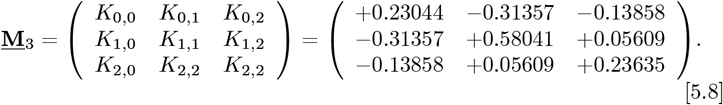

It has a rather small det[<u>**M**</u>_3_] = 1375 value, with an inverse of:

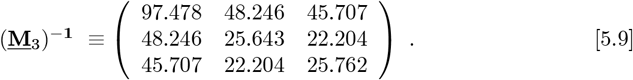

A convolution of *L*_*m*_(*Z*) functions with the measured 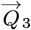dataset vector of Eqs. [4.11a]-[4.11b], along wit h the a bove (<u>**M**</u>_3_)^−**1**^, gives this final 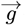:

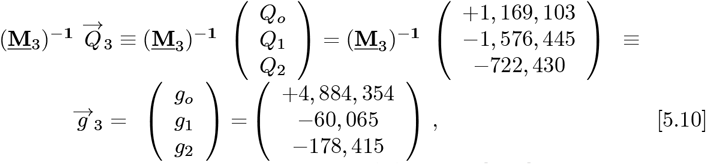

determining the constants needed for *N* (*Z*) in Eq. [2.5a]. The coefficients for *R*(*Z*), which sets the predicted number of daily new CoVID-19 cases, are:

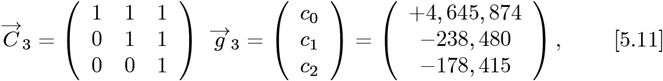

determining the constants needed for *R*(*Z*) in Eq. [2.5b]. Using these {*g*_0_, *g*_1_, *g*_2_} values along with Eq. [2.11] gives:

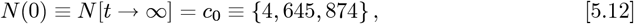

as a new predicted total number of CoVID-19 cases at the pandemic end for the *OFM*, which is a ∼7.54% or 379,026 reduction in the number of cases, compared to the *Initial Model* 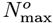 value of Eq. [5.5].

Using Eq. [2.4c] for *Z*[*t*], and substituting the Eq. [5.12] 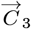 values into Eq. [2.5b] gives *R*(*Z*). The *p*[*t*] in Eq. [2.4a] is derived from *R*(*Z*) using Eq. [2.4b], with the resulting *OFM p*[*t*] plotted in **Figure 4**, using a linear Y-axis, along with the *t*> *t*_*I*_ raw data for the daily new CoVID-19 cases.

**Figure 4:**
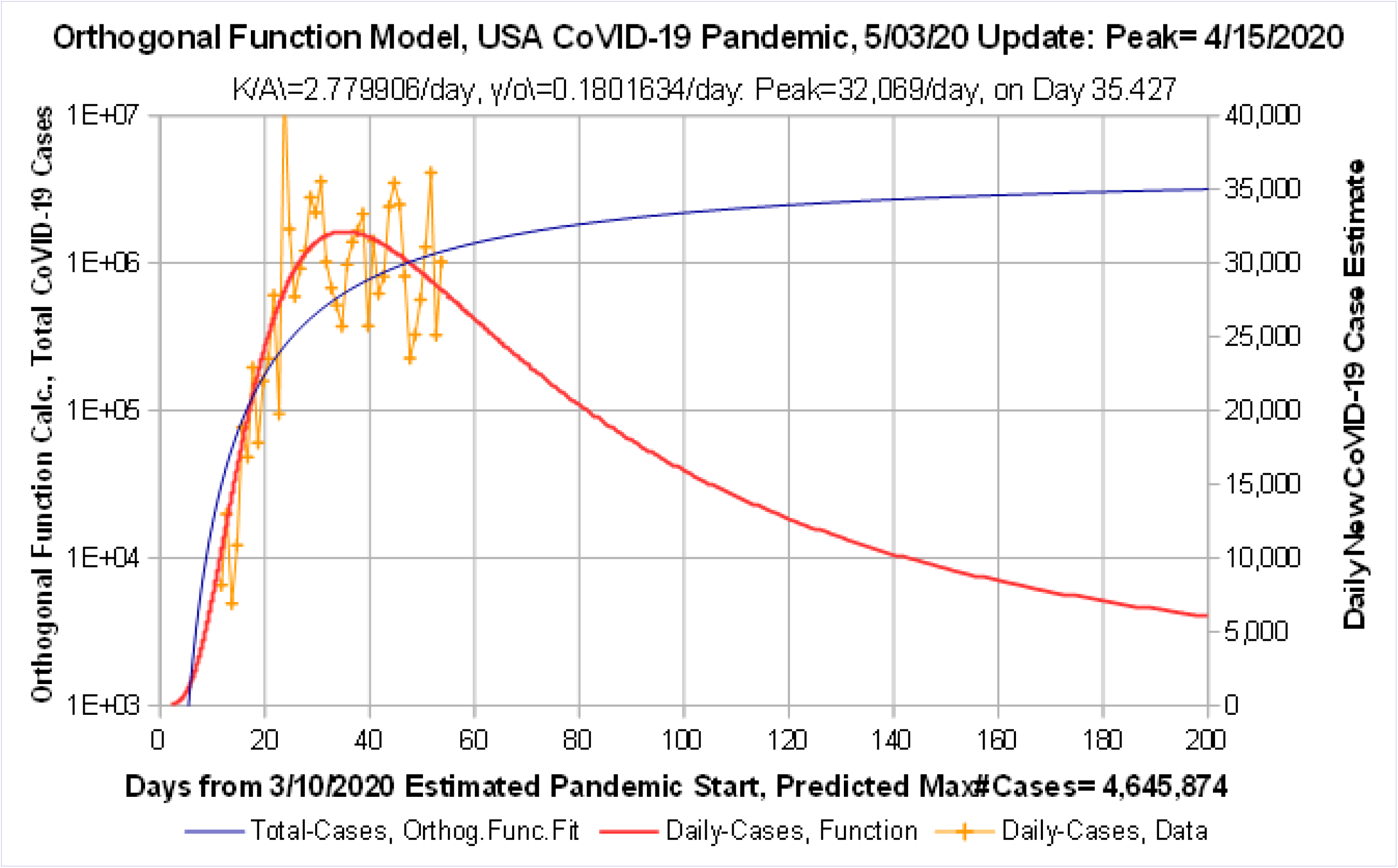
*Orthogonal Function Model*, USA CoVID-19 Projections, data to 5/3/2020. Predicted Number of Daily CoVID-19 Cases has a peak of 32,069 cases/day on 4/15; with 4,645,874 cases total; and ~5,962 new cases/day at Day 200 on 9/26/2020.

Raw data for *t* < *t*_*I*_ was not included in these analyses, because they cover the exponential rise period, prior to *Social Distancing*. Those data are not applicable to estimating *Social Distancing* effects.

However, the **Figure 4** *OFM* provides an extrapolation for those *t* < *t*_*I*_ times, which shows what an exponential rise plus lengthening *doubling times* would have looked like, if both had been operating continuously from the CoVID-19 pandemic start. The companion *N* [*t*] analytic result, plotted using a logarithmic Y-axis, along with the *t*> *t*_*I*_ raw data for the total number of CoVID-19 cases, is show in **Figure 5**.

**Figure 5:**
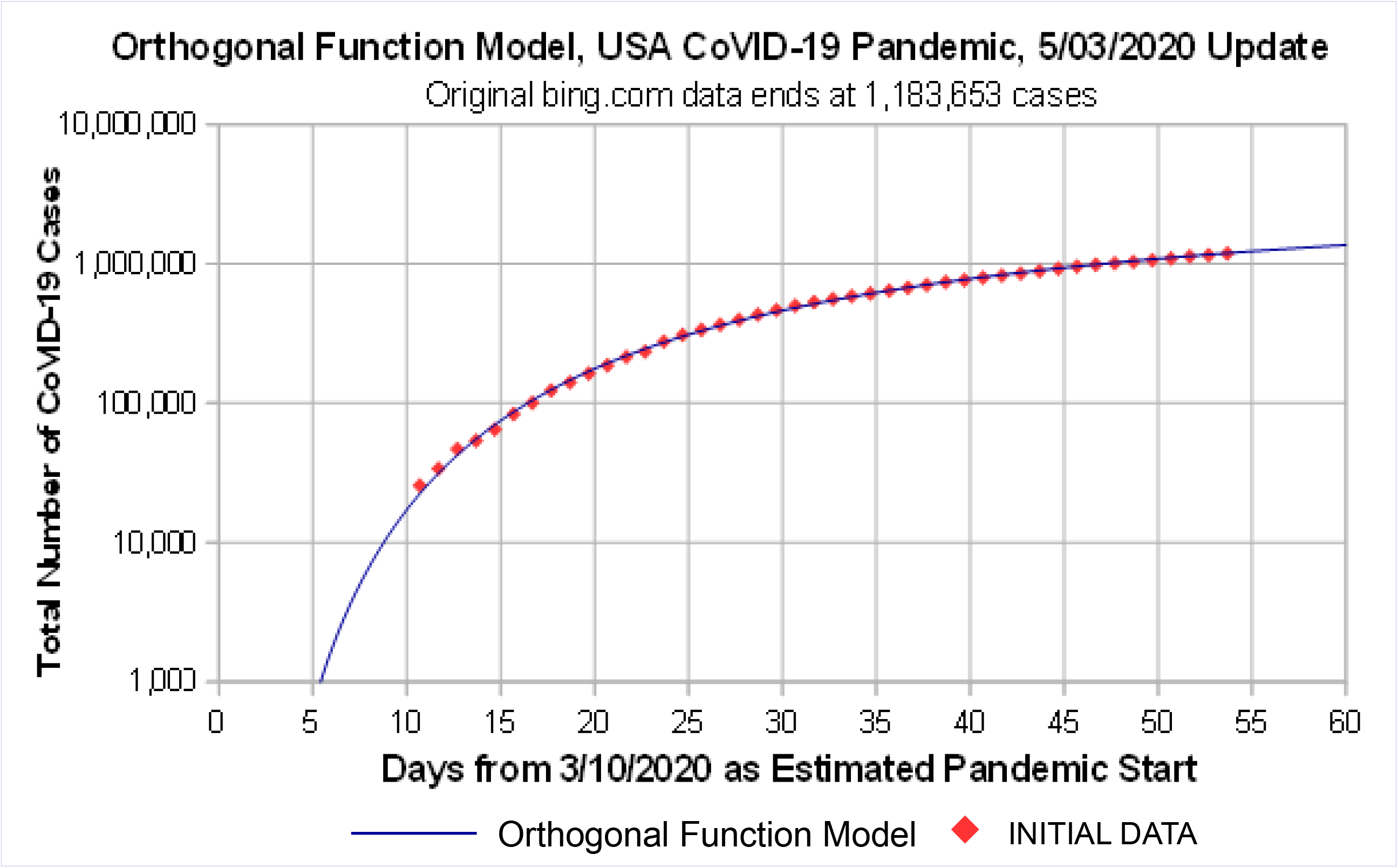
*Orthogonal Function Model*, USA CoVID-19 Projections, data to 5/3/2020. Original *bing.com* data up to 5/3/2020 are shown, prior to their *new reporting method*. *Orthogonal Function* Model matches data a bit better than the *Initial Model*.

Comparing the size and timing of the *p*[*t*] pandemic peak, and its Day 2 value, between the *Initial Model* (**Figs. 2**-**3**) and *OFM* (**Figs. 4**-**5**), gives:

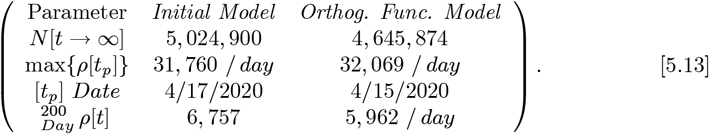

This table shows that the *OFM* predicts fewer total cases (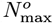 *vs c*_*o*_) and fewer daily new CoVID-19 cases at Day 2, as well as giving an earlier and higher pandemic peak prediction.

While the above analysis used *M*_*F*_ = 2 with Eq. [5.10] 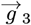 setting the best {*g*_0_, *g*_1_, *g*_2_} values, the *OFM* also provides estimates for the simpler *M*_*F*_ = {0, 1} cases, as outlined by Eqs. [4.9a]-[4.9f]. For *M*_*F*_ = 1, the best two 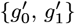 values are gotten by only using {*Q*_0_, *Q*_1_}and an <u>**M**</u>_2_ sub-matrix of <u>**M**</u>_3_. For *M*_*F*_ =, the best 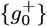 by itself is derived by using {*Q*_0_} and the <u>**M**</u>_1_ sub-matrix. These alternative estimates give:

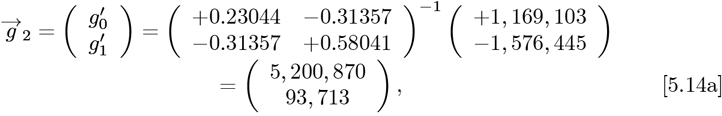

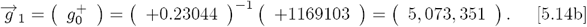

These additional calculations give the following progression of estimates for *N* [*t* → ∞], which is the final n umb er of CoVID-19 cases at the pandemic end:

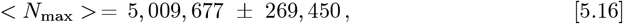

These Eq. [5.15] results show that the *N* [*t* → ∞] projected final number of CoVID-19 cases remains fairly stable, even as the number of data fitting parameters is increased from 0 to 3. The average and 1*σ* standard deviation among these *N*[*t* → ∞] projections is:

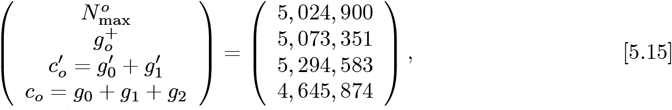

where 1*σ* is ~5:4& of the overall average.

Comparing the results among **Figs. 2**-**5** highlights several items:

A. All *ρ*[*t*] functions have a sharp rise, and a much slower decreasing tail.
B. The overall fit-to-data, as given in **Fig. 3** and **Fig. 5**, shows that the extra parameters in the *OFM* can fit the *ρ*[*t*] shape better.
C. The OFM helps to estimate the uncertainty in the Initial Model, which Eq. [5.16] showed was ~5:4&.
D. These results, taken together, exhibit only a relatively small change in the *N*[*t* → ∞] limits. Thus, the *Initial Model* function captures much of the progression to pandemic shutoff.

The *ρ*[*t*] tail may still differ from these predictions, due to factors such as:

i. The CoVID-19 dynamics may change in the long-term low *ρ*[*t*] regime;
ii. A “second wave” or multiple waves of *ρ*[*t*] rise and fall may occur; both of which are beyond the scope of this CoVID-19 pandemic modeling;
iii. Using just an exponential rise at the CoVID-19 pandemic start, plus lengthening *doubling times*, may limit how much mitigation can be easily modeled using only a few adjustable parameters.

**Figure 4** provides some evidence for the above (iii) possibility. While lengthening the *doubling time* enables pandemic shutoff in the long time *dilute pandemic* limit; **Figure 4** also shows that this model tends to approach final pandemic shutoff rather slowly.

## 6 USA Data: The *bing.com* Change

This analysis of the *bing com* USA data begins at mid-March 2020, when mandatory *Mitigation Measures* were instituted. However, in early-May, *bing com* changed their entire database, revising all numerical values back to the start of their reporting history.

The revised *bing com* USA data from mid-March through early-June is analyzed next, which had these values:

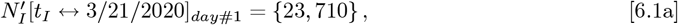

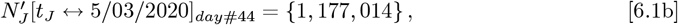

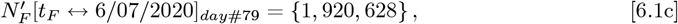

covering 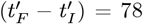 *days*, as compared to the original *bing com* data, which was used in above analyses, and only spanned (*t*_*F*_ − *t*_*I*_) = 43 *days*. The Eqs. [6.1a]-[6.1b] revised {*day* #1, *day* #44} values are {∼7082%, ∼ 56%} lower than the original Eqs. [5.1a]-[5.1b] data.

Applying the *Initial Model* to this revised dataset gives:

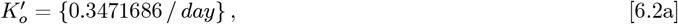

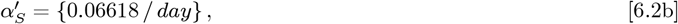

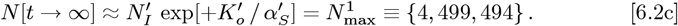

Using Eq. [3.3b] gives these 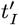 and 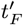 results:

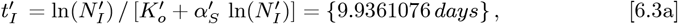

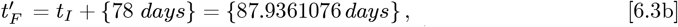

for use in the *OFM*. The Eq. [6.2c] calculated 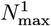 value is ∼10.456% lower than the prior 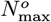 of Eq. [5.5]. Since the *Initial Model* uses an *rms* best fit on logarithmic axes for *N* [*t*], it emphasizes differences at low *N* [*t*] values, where the revised *bing com* data changes were larger. Thus, some of the ∼10.456% change in 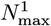 may be due to the revised *bing com* data, but the longer 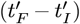 data interval also contributes to modifying the 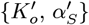 values.

The *Initial Model* datafit for the revised USA data is shown in **Figures 6**-**7**, and is a better datafit than the *Initial Model* results of **Figures 2**-**3**. Comparing the *OFM* result of Eq. [5.12], which gave *N* [*t*→ ∞] = {4, 645, 874}, to the *Initial Model* result of Eq. [6.2c] shows that they differ by just ∼3025%.

**Figure 6:**
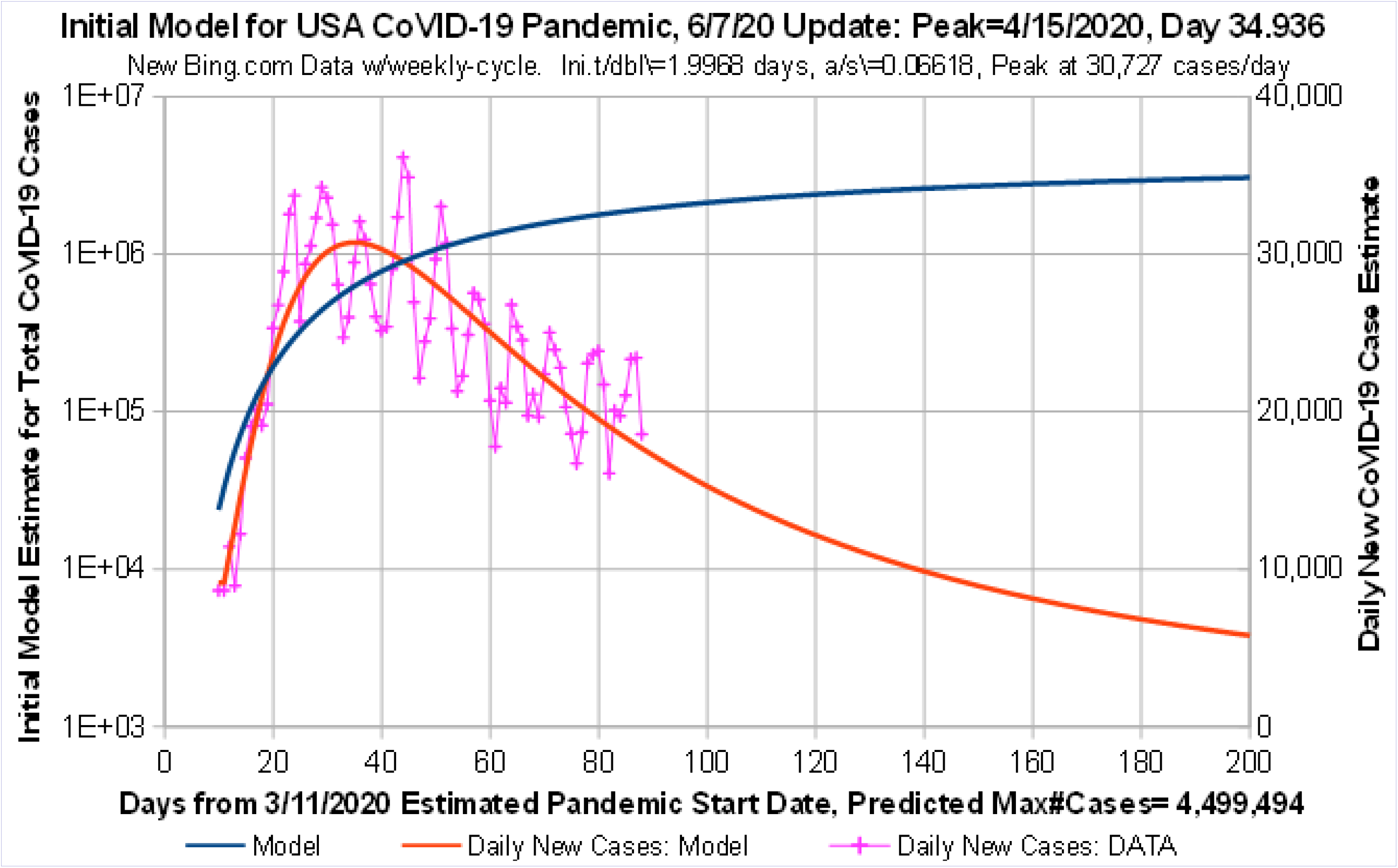
*Initial Model* for USA CoVID-19 Projections vs data up to 6/7/2020. Predicted Number of Daily CoVID-19 Cases has a peak of 30,727 cases/day on 4/15; with 4,499,494 cases total; and ~5,783 new cases/day at Day 200 on 9/27/2020.

Next, the *OFM* is applied to further refine this *Initial Model* prediction. Those results are shown in **Figure 8** and **Figure 9**, which were derived as follows. First, the Eqs. [2.1a]-[2.1d] time-shift was done:

**Figure 7:**
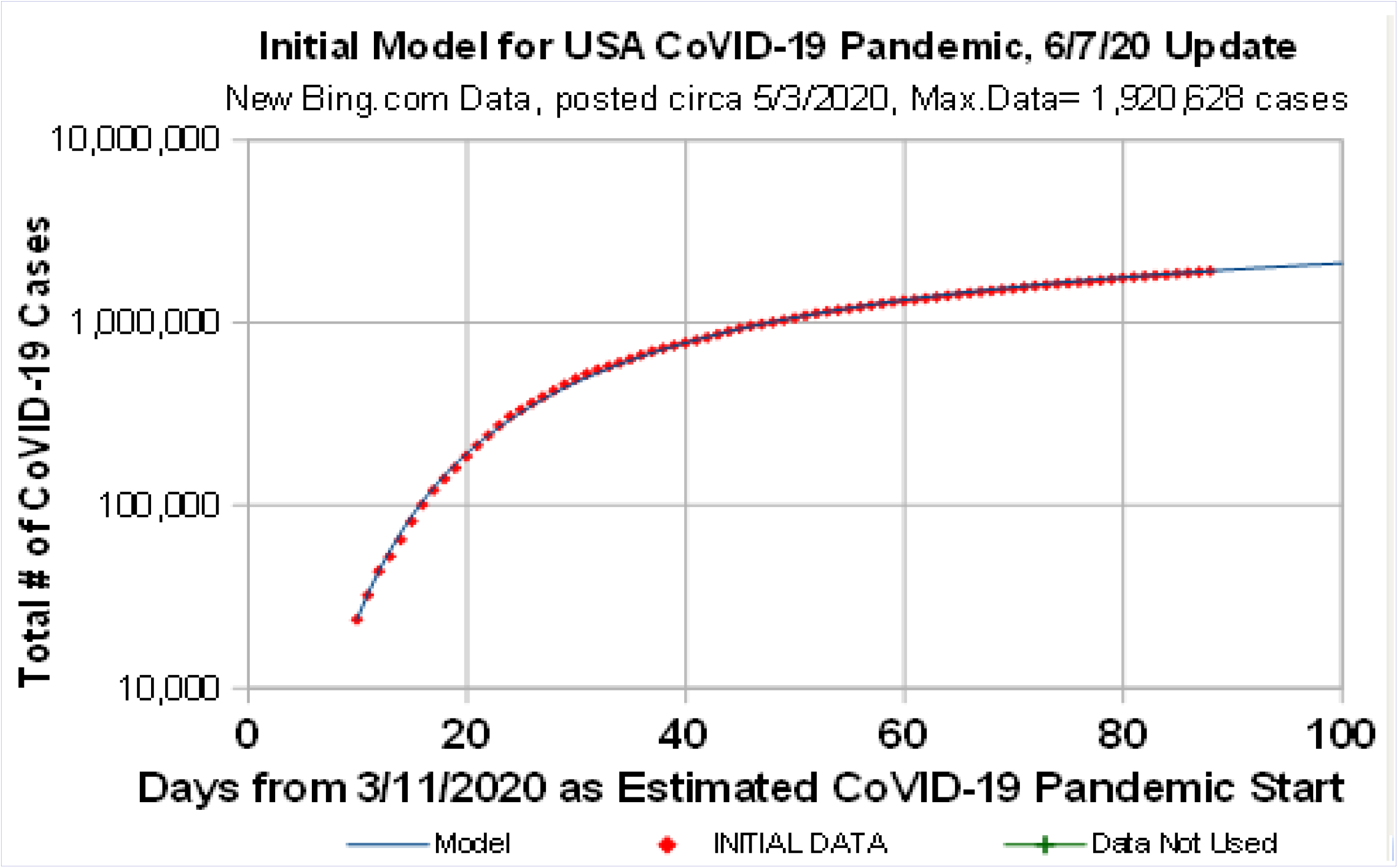
*Initial Model* for USA CoVID-19 Projections vs data up to 6/7/2020. Revised *bing.com* data, circa 5/3/2020, changed all values back to the pandemic start. *Initial Model* appears to be a good data.t by itself.

**Figure 8:**
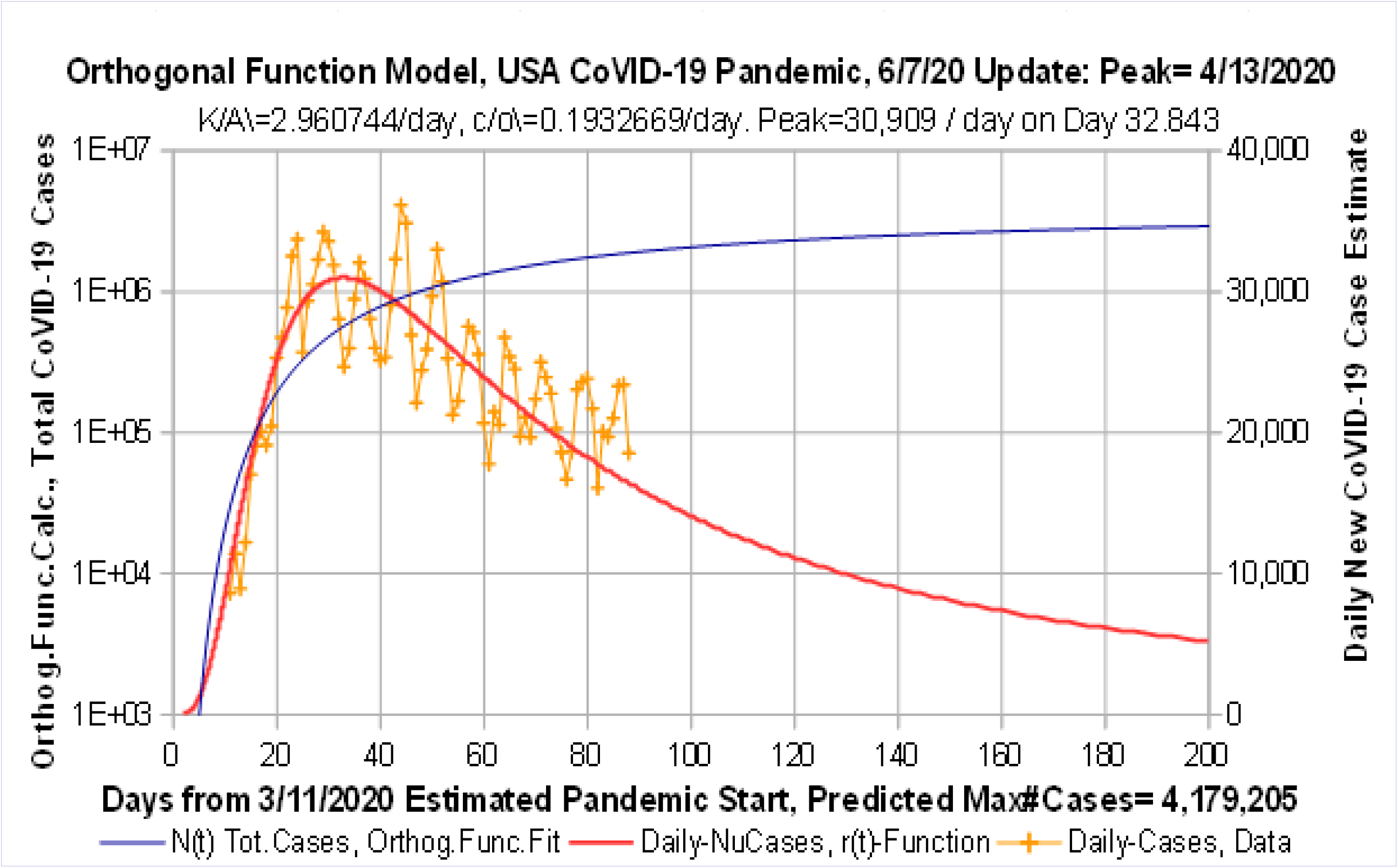
*Orthogonal Function Model*, USA CoVID-19 Projections, data to 6/7/2020. Revised *bing.com* data; daily# of CoVID-19 Cases Peak at 30,909 cases/day on 4/13/2020; with 4,179,205 cases total; and ~5,140 new cases/day at Day 200 on 9/27/2020.

**Figure 9:**
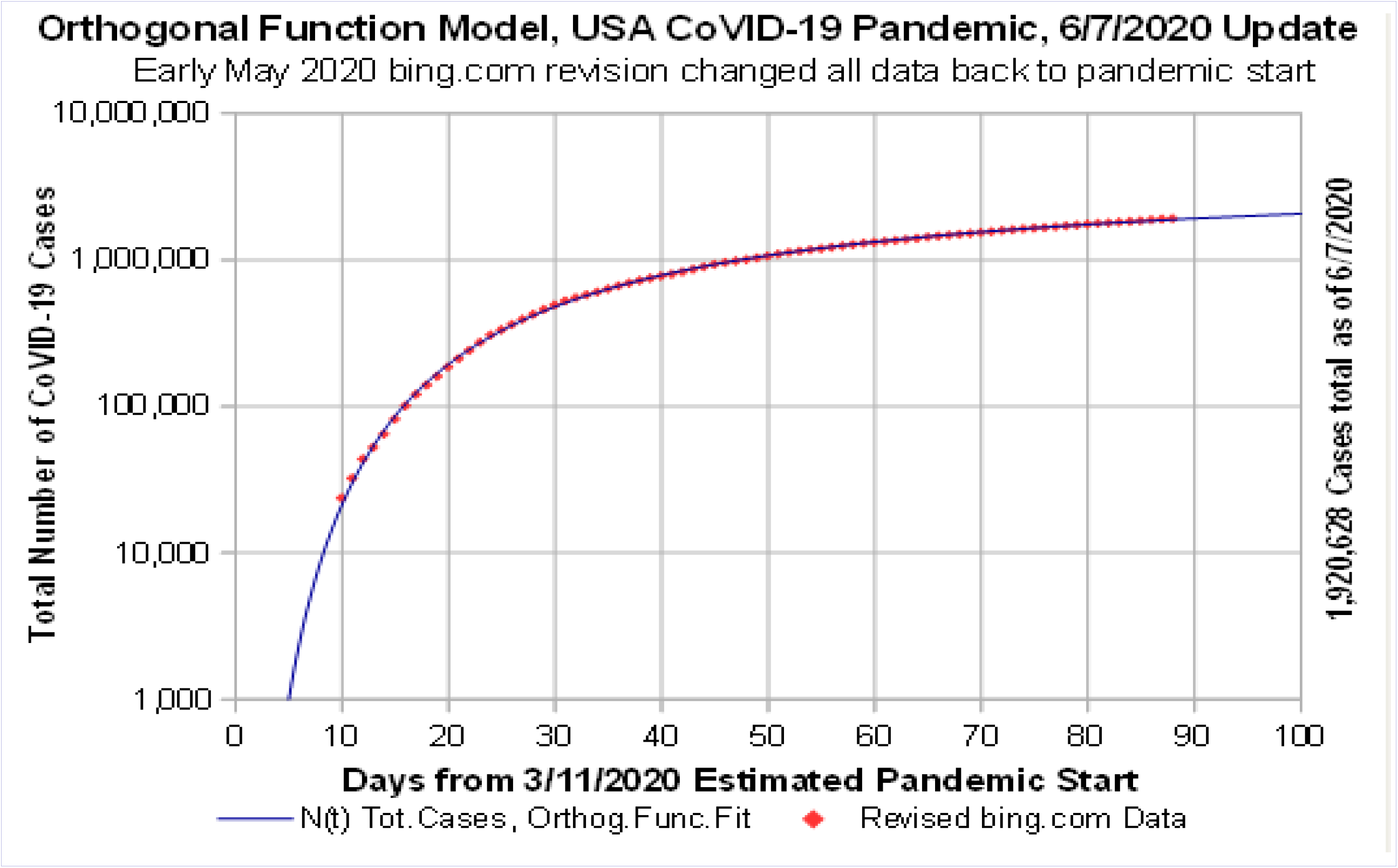
*Orthogonal Function Model*, USA CoVID-19 Projections, data to 6/7/2020. Revised *bing.com* data, posted circa 5/3/2020, changed values back to the pandemic start. *Orthogonal Function Model* matches the data a bit better than the *Initial Model*.

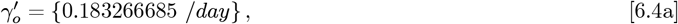

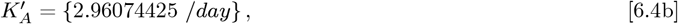

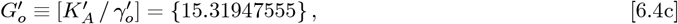

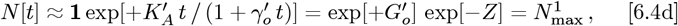

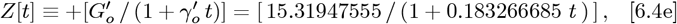

for this dataset. Next, using Eqs. [6.3a]-[6.3b] for 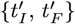 gives:

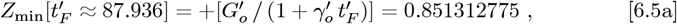

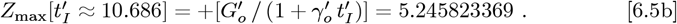

The <u>**M**</u>_3_ matrix of *K*_*m*,*n*_ entr ies, a s set by the {*Z*_min_, *Z*_max_} values, is:

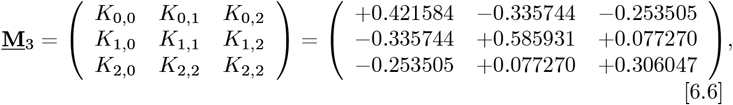

and it has an inverse of:

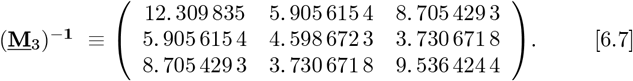

The 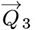 for this dataset gives this up dated 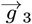– vector:

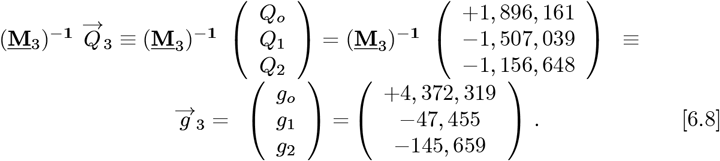

where *c*_0_ = (*g*_0_ + *g*_1_ + *g*_2_) = (4,0179, 205) = *N* [*t* →∞] is the new *OFM* predicted total number of CoVID-19 cases, which is down from the *Initial Model* value of 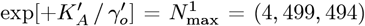 from Eq. [6.2c]. This ∼7012% reduction is similar to the ∼7042% change between Eq. [5.12] and Eq. [5.5]. A similar analysis for 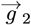 and 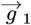, using Eqs. [4.9a]- [4.9f], gives this summary:

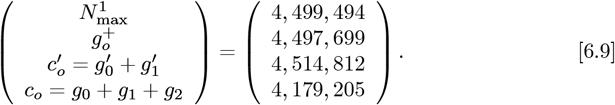

The *N* [*t* → ∞] projected final number of CoVID-19 cases in Eq. [6.9] remains fairly stable, even as the number of data fitting parameters is increased from 0 to 3. This result is similar to the Eq. [5.15] analysis of the original *bing com* data, which spanned only (*t*_*F*_ − *t*_*I*_) = 43 *days*. The average and 1*α* standard deviation among these Eq. [6.9] calculations for *N* [*t*→ ∞] is:

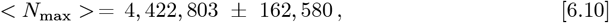

where 1*σ* is ∼3.68% of the overall average. It is somewhat lower than the ∼5014% value of Eq. [5.16]. Thus, having 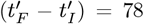 *days* of data for analysis reduces the overall uncertainty in these projections.

Comparing Eq. [6.9] to Eq. [5.15] also shows the following trends. The 1-term calculations, using either {*N*_max_} or just 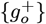 by itself, give similar results. The 2-term calculations, using 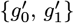 gives ≲ 10 % higher results, while using {*g*_0_, *g*_1_, *g*_2_} gives ≲ 1 % lower results. This oscillation around the average value of Eq. [6.10] shows that the *Initial Model* of Eq. [2.1a] and Eq. [6.4d] capture much of how *Social Distancing* enables pandemic shutoff.

Comparing the **Fig. 6** *Initial Model* and the **Fig. 8** *OFM* for the pandemic peak size, timing, and Day 2 values gives:

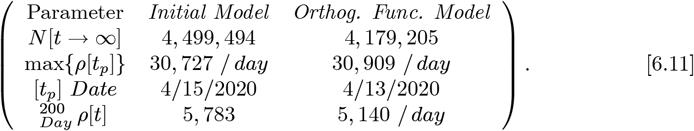

The *p*[*t* → ∞] at 2-days nearly scales with *N* [*t* → ∞], while the *OFM* predicts a higher and earlier *ρ*[*t*] pandemic peak. Comparing the revised *bing com* 78-day dataset up through 6/7/2020 of Eq. [6.11], to the original *bing com* 43-day dataset up through 5/3/2020 of Eq. [5.13] gives:

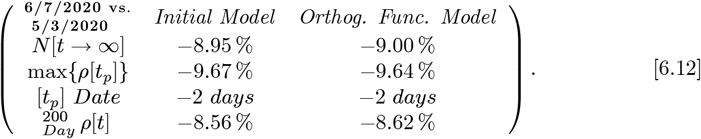

Both the *Initial Model* and the *OFM* found a comparable amount of change between the two datasets; likely due to the revised *bing com* values being lower, along with the larger dataset enabling increased modeling precision.

The *Initial Model* and the *OFM* also provide self-consistent CoVID-19 predictions over the two different time periods. Each model held its predictive power to within < 10 % for over a month 43 *days vs* 78 *days*, without needing recalculations or parameter value changes, which provides a strong data-driven validation of the potential utility of these models. When the *Initial Model* is a somewhat good fit, this *Orthogonal Function Model* provides even better fits.

## 7 Italy: Revised *bing.com* Data Analysis

This Italy analysis uses data beginning on Feb. 23, 2020, from the revised *bing com* CoVID-19 database^**9**^, which has these values:

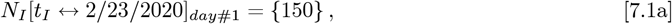

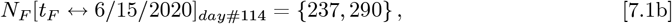

with (*t*_*F*_ − *t*_*I*_) = 113 *days*. The number of daily new CoVID-19 cases shows a sharp post-peak decrease for Italy, in contrast the the above USA data. That sharp decrease provides a near-worst case test for the *OFM*. The *Initial Model* best fit on a logarithmic Y-axis, gives these initial parameters:

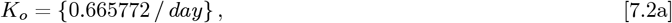

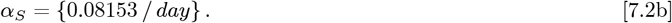

Using Eq. [3.3b] for *t*_*I*_ and *t*_*F*_ gives:

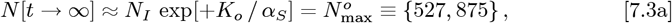

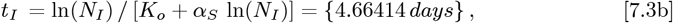

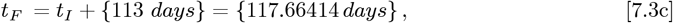

for use in the *OFM*. The revised *bing com* Italy data and the *Initial Model* datafit are shown in **Figure 1** and its inset. The *Initial Model* is not a good fit due to the high curvature of the data on the logarithmic Y-axis, which is similar to our previous^**1**^ results for Italy. The *OFM* is applied next.

Using Eqs. [3.5a]-[3.5b] sets these {*γ*_*o*_, *K*_*A*_, *G*_0_} values:

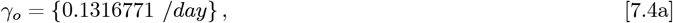

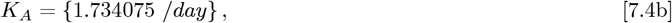

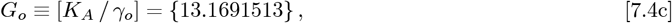

where Eq. [3.1c] also gives:

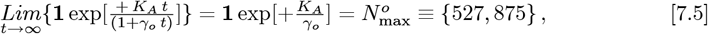

which matches Eq. [7.3a], as it should. Then:

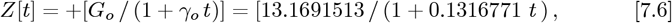

defines *Z* for the *OFM*, where:

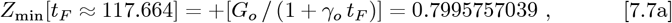

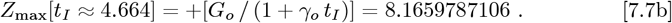

The resultant symmetric matrix <u>**M**</u>_3_ of *K*_*m,n*_ entries is:

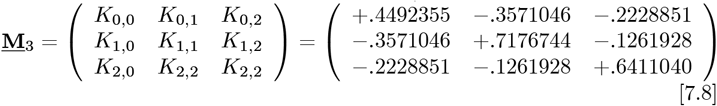

It has an (<u>**M**</u>_3_) inverse of:

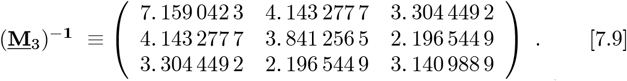

A convolution of *L*_*m*_(*Z*) functions with the measured data sets 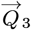 using Eqs. [4.11a]-[4.11b], with (<u>**M**</u>_3_)^−**1**^ of Eq. [4.5c] giving this final 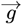 vector:

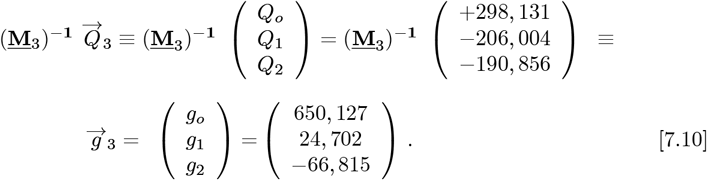

The coefficients for *R*(*Z*), which set the predicted number of daily new CoVID-19 cases for the *OFM*, are given by:

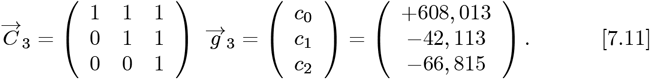

Using these *g*_0_, *g, g* values along with Eq. [2.11] gives:

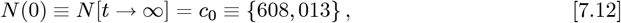

as a new predicted total number of CoVID-19 cases at the pandemic end. It is a ∼15.18% or 800,0138 increase in number of cases, compared to the *Initial Model* 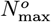 value of Eq. [7.3a].

A graph of *ρ*[*t*] for the predicted number of daily new CoVID-19 cases is shown in **Figure 11**, using Eqs. [2.4b] and [2.5b]. For this fast pandemic shutoff case, the *OFM* improvement over the *Initial Model* is not large. hen the initial [exp(−*Z*)] function is not a good fit, which is likely for quicker pandemic shutoffs, a lot of terms, beyond the *M*_*F*_ = 2 value used here, are needed in Eq. [2.5a] for a good fit. An alternative method for choosing the initial [exp(−*Z*)] function is examined next, to see if additional improvements result for that case.

**Figure 10:**
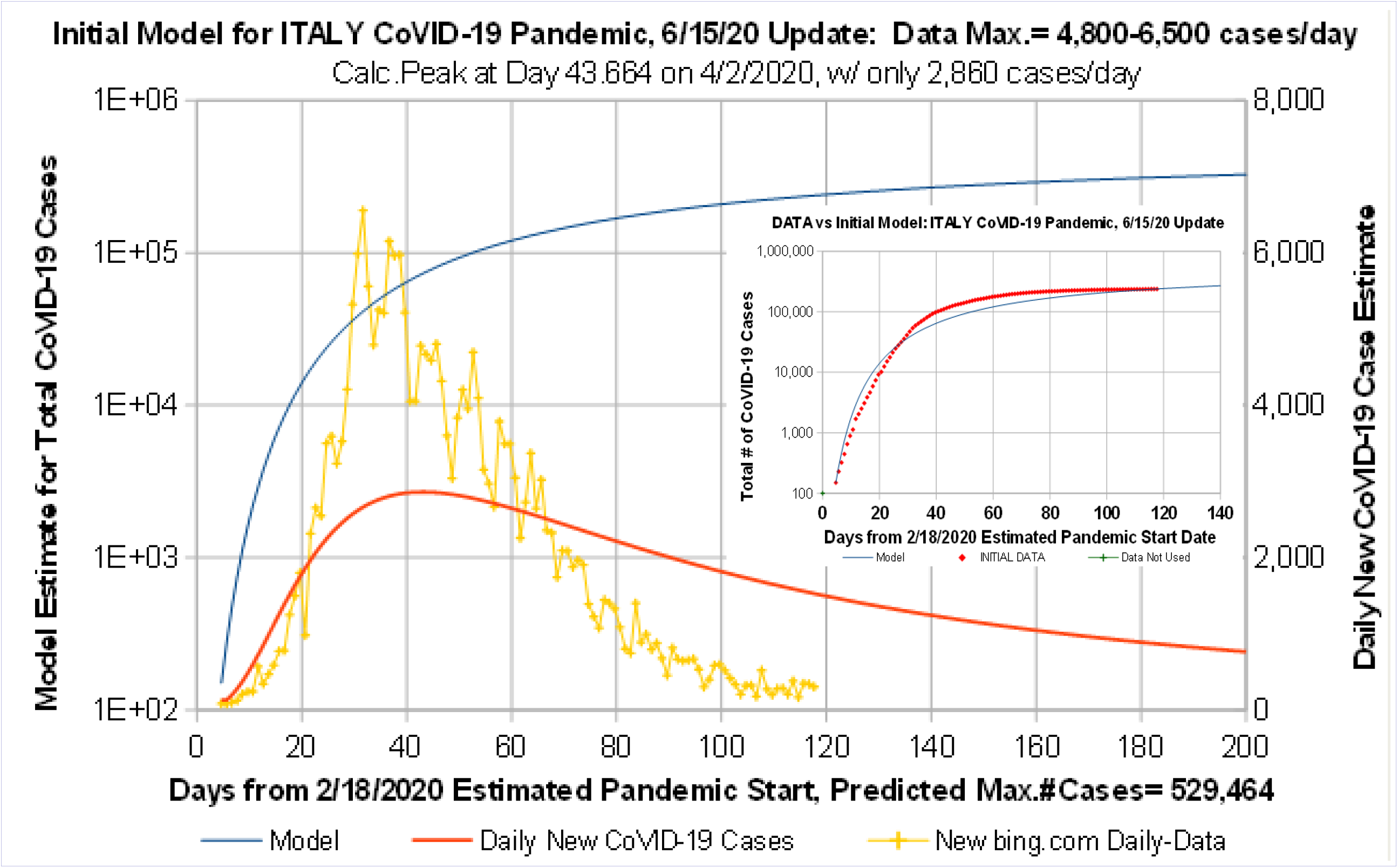
*Initial Model* for ITALY CoVID-19 Projections vs data up to 6/15/2020. *Initial Model* matches Total Number of Cases at data start and data end, but best fit using a logarithmic Y-axis does not give a good fit for Predicted Number of Daily CoVID-19 cases.

**Figure 11:**
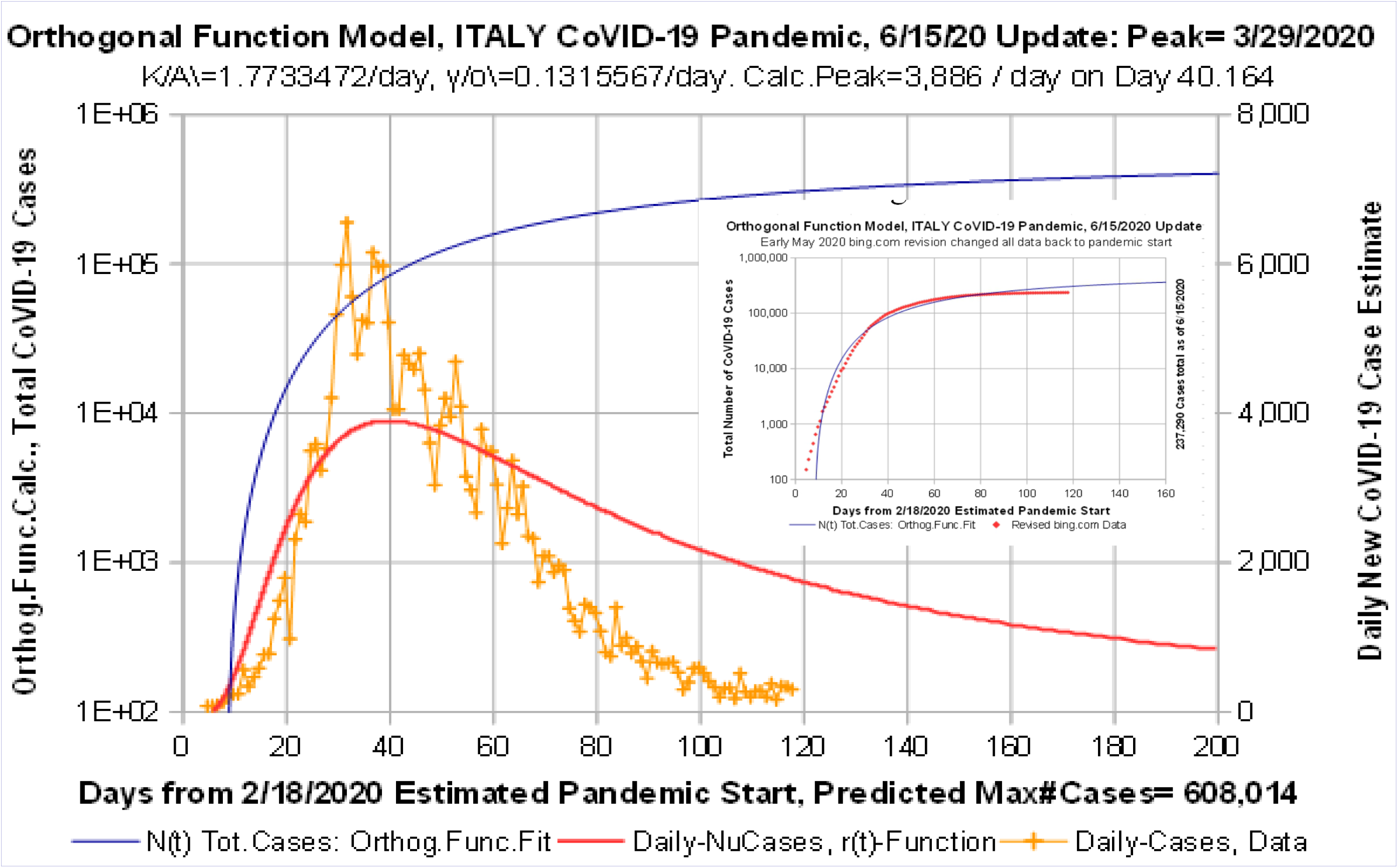
*Orthogonal Function Model* for ITALY CoVID-19 data up to 6/15/2020. *Orthogonal Function Model* gives improved datafit, but 3-terms in orthogonal function series is insufficient to accurately predict a rapidly decreasing Number of Daily CoVID-19 cases.

## 8 Italy: An Alternative Starting Function

There is a wide latitude in the choice of an initial [exp(−*Z*)] function for the Eqs. [2.5a]-[2.5b] orthogonal function expansions. However, when the *Initial Model* is not a good fit, the common practice of minimizing *rms* error using a logarithmic Y-axis for the *Initial Model* may not be optimal, since the *Orthogonal Function Model fOFMj* creates best fits using a linear Y-axis.

Minimizing the *rms* error between the *Initial Model* and data using a linear Y-axis is done to provide an alternative [exp(−*Z*)] function. This alternative starting point gives these parameter values, replacing Eqs. [7.2b]-[7.2c]:

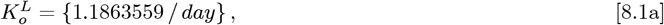

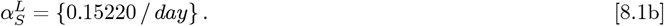

The *N* (*t*_*I*_) = *N*_*I*_ and *N* (*t*_*F*_) = *N*_*F*_ values are still needed to properly set the above 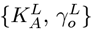 values for *Z*[*t*]. Using Eq. [3.3b] for *t*_*I*_ and *t*_*F*_ gives:

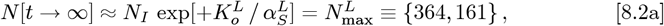

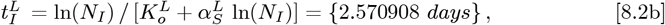

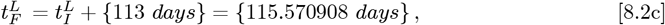

for use in the *OFM*, while still using this linear Y-axis initial fit. **Figure 12** and its inset show how this alternative *Initial Model* compares to the Italy CoVID-19 data. Using Eqs. [3.5a]-[3.5b] sets these new 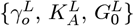 values:

**Figure 12:**
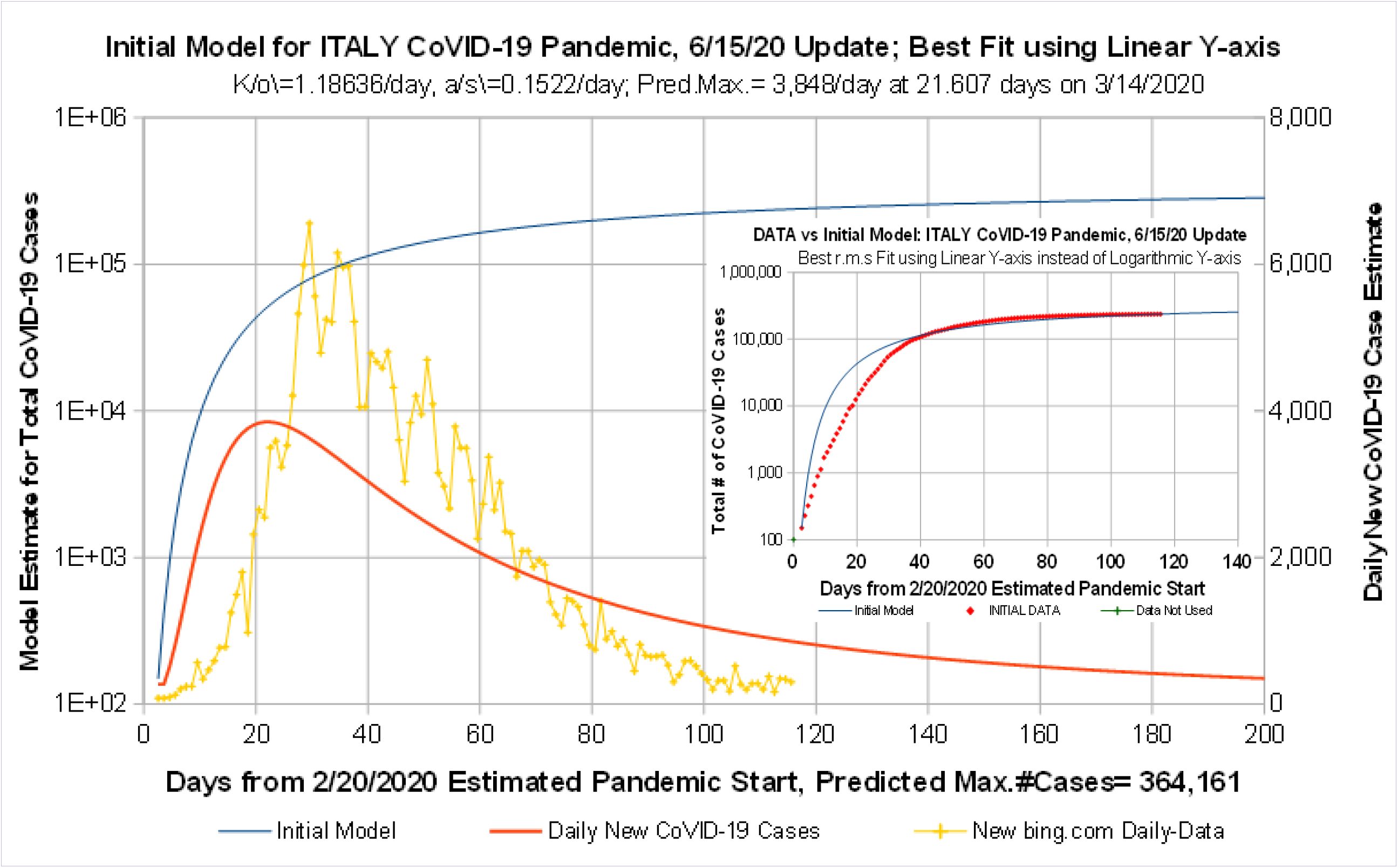
*Initial Model* re-do, ITALY CoVID-19 data to 6/15/2020. New starting point is a best fit function on a linear Y-axis, instead of having a best fit using a logarithmic Y-axis. Alternative method may allow a few-term orthogonal function series to better match the data.

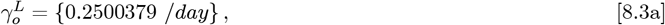

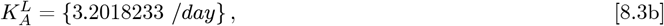

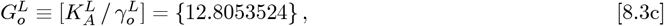

where Eq. [3.1c] also gives:

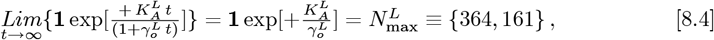

which matches Eq. [8.2a], as it should. Then:

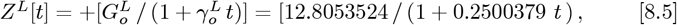

defines *Z*^*L*^ for this alternative fit analysis, where:

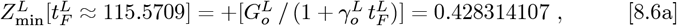

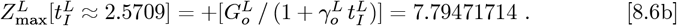

The resultant symmetric matrix **<u>**M**</u>**_3_ of *K*_*m,n*_ entries is:

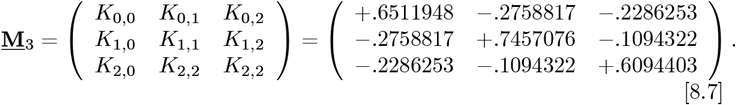

It has an (**<u>**M**</u>**_3_) inverse of:

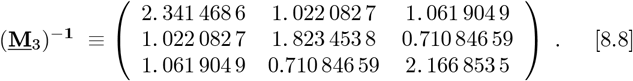

A convolution of *L*_*m*_(*Z*^*L*^) functions with the measured data sets 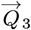_3_ using Eqs. [4.11a]-[4.11b], with (**<u>**M**</u>**_3_)^-**1**^ of Eq. [4.5c] giving this final 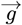– vector:

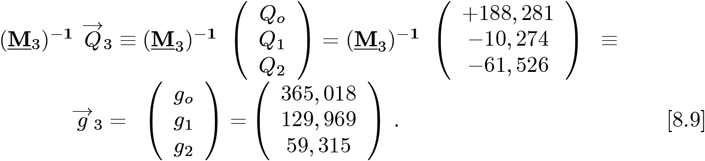

The coefficients for *R*(*Z*), which set the predicted number of daily new CoVID-19 cases for the *OFM*, are given by:

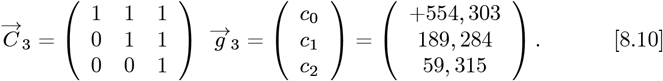

Using these {*g*_0_, *g*_1_, *g*_2_} values along with Eq. [2.11] gives:

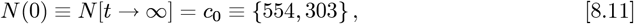

as a new predicted total number of CoVID-19 cases at the pandemic end. It is a ∼5 % or 26, 428 increase in number of cases, compared to the *Initial Model* 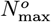 value of Eq. [7.3a]. A graph of *p*[*t*] for the predicted number of daily new CoVID-19 cases is shown in ****Figure 13****, using Eqs. [2.4b] and [2.5b]. A tabulated summary for all of these Italy calculations is:

**Figure 13:**
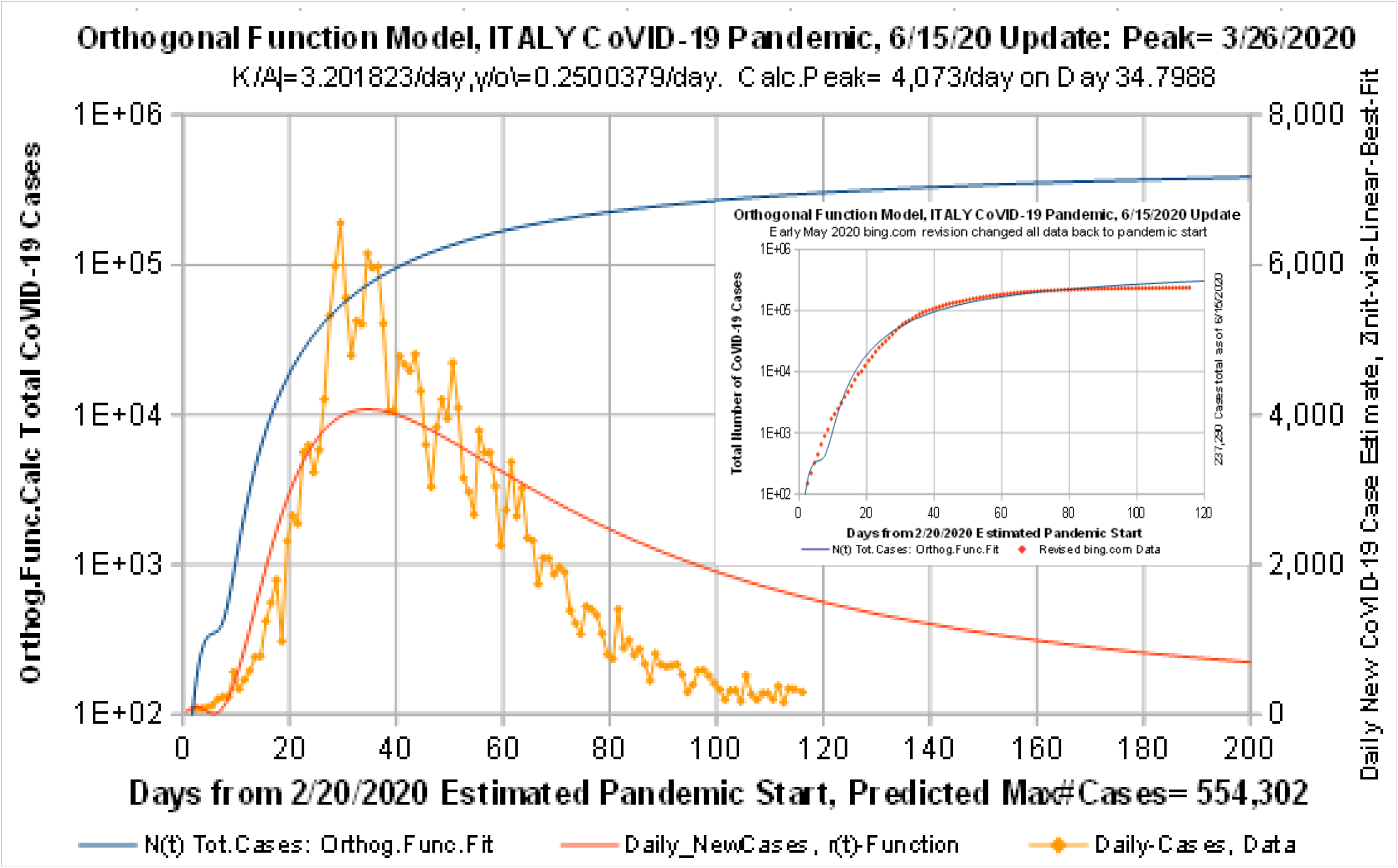
*Orthogonal Function Model* re-do, ITALY CoVID-19 data to 6/15/2020. *Orthogonal Function Model* re-do using linear Y-axis gives a slightly better small-series fit. Other *Social Distancing* impacts likely exist besides just lengthening pandemic *doubling times*.

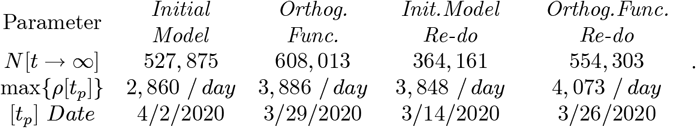

The *Initial Model* shapes for *ρ* [*t*] were very different, depending on whether that initial datafit was performed by minimizing *rms* error using a logarithmic Y-axis (****Figure 10****) or a linear Y-axis (****Figure 12****, *Initial Model Re-do*) as expected. However, comparing the two *OFM* (****Figure 11**** vs ****Figure 13****) calculations, shows that their overall *ρ* [*t*] shapes are quite similar.

While the max {*ρ* [*t*_*p*_]} calculated pandemic peaks generally increase, they are all below the data near-peak values of ∼4, 800− 6, 5 cases/day shown in **Figs. 10**-**13**. Thus, for quick pandemic shutoffs, the *Initial Model* [exp(−*Z*)] function is less important than needing more *M*_*F*_ terms. hen the *Initial Model* is not a good fit, the *OFM* only gives limited improvements for *M*_*F*_ = 2.

## 9 Summary and Conclusions

The early stages of the CoVID-19 coronavirus pandemic began with a nearly exponential rise in the number of infections with time. Let *N* [*t*] be the total number of CoVID-19 cases vs time. Our *Initial Model*^**1**^ used this basic function:

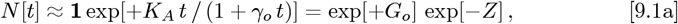

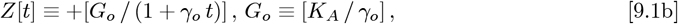

to model *Social Distancing* effects by progressively lengthening the *doubling time* for the pandemic growth. The *γ*_*o*_ = limit of Eq. [9.1a] corresponds to a purely exponential rise. This *Initial Model* enables calculation of a pandemic shutoff with only a small fraction of the total population becoming infected (*“dilute pandemic”*).

To allow more data fitting parameters than just { *K*_*A*_, *γ*_*o*_}, an *Orthogonal Function Model* |*OFM*| was developed, using these orthogonal function series:

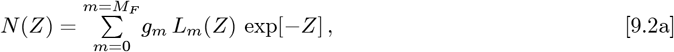

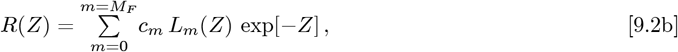

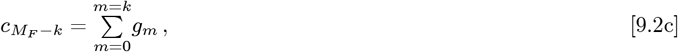

where *N* [*t*] = *N* (*Z*[*t*]). The {*g*_*m*_; *m* = (0, +*M*_*F*_)} are a set of constants that are determined from each dataset. Using exp [−*Z*] as a weighting function, with *L*_*m*_(−*Z*) as an orthonormal function set on the *Z* ={ 0^+^, ∞^−^} interval, the choice of *L*_*m*_(*Z*) becomes unique. They are the *Laguerre Polynomials*, with several important properties given in Eqs. [2.6a]-[2.7e].

The expected number of daily new CoVID-19 cases, *p*[*t*], is given by:

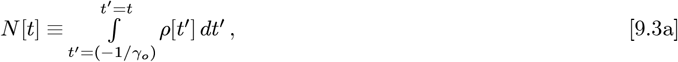

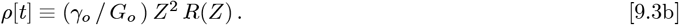

For a wide range of *N* (*Z*) data, larger *M*_*F*_ and more {*L*_*m*_(*Z*); *m* = (0, +*M*_*F*_)}terms gives progressively better matches to almost any *arbitrary function*, enabling improved data fitting for a variety of *N* [*t*] and *ρ* [*t*] shapes.

Methods are developed here to derive {*K*_*A*_, *γ*_*o*_}, and determine the {*g*_*m*_; *m* = (0, +*M*_*F*_)} and { *c*_*m*_; *m* = (0, +*M*_*F*_)}constants from any given *N* [*t*] dataset. Whereas our *Initial Model* was an *M*_*F*_ = case, the *M*_*F*_ = 2 case was used here for data analysis, as an *OFM* example.

These methods were applied to the CoVID-19 pandemic data for the USA. Analysis results using the original *bing com* up data to~5/3/2020 are given in **Figures 2**-**5** and Eq. [5.13]. During early-May, *bing com* revised their entire database, all the way back to their earliest values. This revised USA *bing com* data, which included an extended time period into June 2020, was also analyzed, with results given in **Figures 6**-**9** and Eq. [6.11].

For the USA, the *Initial Model* and *OFM* results differed by only ∼1 %, showing that the *Initial Model* was a somewhat good fit, while the *OFM* is a better fit. Comparing our calculations using the 43-day 5/3/2020 original *bing com* dataset to the 78-day 6/7/2020 revised *bing com* dataset, showed that our early-May USA projections predicted the June data to within ≲10 % for the same model. Thus, both models provided self-consistent CoVID-19 projections, holding their predictive power for over a month { 43 *days vs* 78 *days*}, without recalculations or parameter value changes.

The Italy CoVID-19 pandemic data was studied next, as a worst-case test of the *OFM*. The post-May 2020 revised *bing.com* database was used, with results presented in **Figures 10**-**13**. Italy had a much sharper CoVID-19 pandemic shutoff for *ρ* [*t*] compared to the USA. hile the *OFM* can give substantial improvements, here *M*_*F*_ = 2 does not provide enough extra parameters, to convert an *Initial Model* result that was not a good fit, into a substantially better fit. A larger *M*_*F*_ and additional orthogonal function terms are needed.

Even then, the long-term tail can be inaccurate, since both the *Initial Model* and the *OFM* extension have natural *ρ*[*t*] asymptotic limits of *ρ*[*t*]∼[1/*t*^**1**^]. Larger *M*_*F*_ values could allow multiple terms to cancel, but a polynomial-like tail of *ρ*[*t*]∼[1/*t*^*P*^], with *P* ≥ 2, would likely remain, making it difficult to estimate the functional form of the CoVID-19 tail for quick pandemic shutoffs.

Overall, both the *Initial Model* and this *Orthogonal Function Model* show how progressively lengthening the pandemic *doubling time* enables CoVID-19 pandemic shutoff, even in the *dilute pandemic* limit. However, there may a natural limit to how fast this one mitigation factor can achieve pandemic shutoff. For cases like Italy, other *Social Distancing* factors may be operating that enable and enhance quick CoVID-19 pandemic shutoff, which are **not** effectively being modeled by just lengthening the pandemic *doubling times*.

## Data Availability

All data used is in the public domain or was maintained by bing.com.

